# Radiation doses to cardiac substructures predict elevation in high-sensitivity cardiac troponin T (hs-cTnT) levels in radiotherapy for lung cancer

**DOI:** 10.1101/2025.09.09.25335431

**Authors:** Xinru Chen, Xiaodong Zhang, Ting Xu, Radhe Mohan, Ruitao Lin, Mei Chen, Rachel C. Maguire, Yao Zhao, Efstratios Koutroumpakis, Nicolas L. Palaskas, Anita Deswal, Ali Ajdari, Joshua S. Niedzielski, Sanjay S. Shete, Laurence E. Court, Jinzhong Yang, Zhongxing Liao

**Affiliations:** Department of Radiation Physics, Division of Radiation Oncology, The University of Texas MD Anderson Cancer Center, Houston, Texas, USA; The University of Texas MD Anderson Cancer Center UTHealth Houston Graduate School of Biomedical Sciences, Houston, Texas, USA; Department of Radiation Oncology, Division of Radiation Oncology, The University of Texas MD Anderson Cancer Center, Houston, Texas, USA; Department of Biostatistics, Division of Basic Science Research, The University of Texas MD Anderson Cancer Center, Houston, Texas, USA; Department of Cardiology, Division of Internal Medicine, The University of Texas MD Anderson Cancer Center, Houston, Texas, USA; Department of Radiation Oncology, Mass General Research Institute, Boston, Massachusetts; Department of Thoracic Radiation Oncology, Division of Radiation Oncology, The University of Texas MD Anderson Cancer Center, Houston, Texas, USA

**Keywords:** cardiotoxicity, logistic regression, radiomics, outcome prediction

## Abstract

**Purpose:** Cardiotoxicity is a major concern for patients undergoing thoracic radiotherapy. This study compared the predictive power of radiation dose-volume histogram (DVH) parameters and radiomic/dosiomic features of the whole heart (WH) and cardiac substructures for elevated circulating high-sensitivity cardiac troponin T (hs-cTnT), a biomarker for early detection of cardiac adverse events.

**Methods and Materials:** A retrospective cohort of 160 patients with non-small cell lung cancer (NSCLC) from a completed prospective trial and a prospective cohort of 57 patients with NSCLC enrolled in an ongoing trial were analyzed. The endpoint was hs-cTnT elevation, indicated by increase of ≥5 ng/L from baseline. An in-house auto-segmentation model delineated 19 cardiac substructures. DVH parameters, radiomic, and dosiomic features were extracted from each patient. A 100-iteration Monte Carlo cross-validation (75%/25% split) was conducted within the retrospective cohort to mitigate random split bias. Logistic regression models using different input combinations were compared, with a model using only clinical factors served as the baseline. Key predictive features were identified using permutation importance during training. Models were validated by hold-out in the prospective cohort to evaluate robustness.

Model performance was assessed by area under the receiver operating characteristic curve (AUROC).

**Results:** Incidence of hs-cTnT elevation was 31.9% in the retrospective and 29.8% in the prospective cohort. The substructure DVH model achieved the highest predictive performance in cross-validation (mean AUROC 0.71, 95% CI [0.70, 0.73]) and demonstrated greater robustness in hold-out validation compared to WH-based models (AUROC 0.60 vs. ≤0.51). Feature analysis identified the left anterior descending coronary artery V20Gy as the most dominant predictor, with cut-off values ranging from 0.2%-5% using various indices.

**Conclusions:** Cardiac substructure DVH parameters have superior predictive power and robustness over WH variables for predicting hs-cTnT elevation in NSCLC radiotherapy, emphasizing the need of using cardiac substructures for cardiotoxicity risk assessment.

## INTRODUCTION

Cardiotoxicity has long been recognized as a major complication for patients receiving thoracic radiotherapy, with substantial evidence linking thoracic irradiation to increased cardiac risk.^1–4^ Cardiac diseases accounted for 5.3% of total deaths in a disease-specific mortality analysis of more than 270,000 lung cancer patients, ranking second only to tumor-related mortality.^5^ Moreover, radiation-related cardiovascular mortality may be underestimated owing to frequent cardiac-related hospitalizations and numerous sudden deaths without documented cancer recurrence.^6^ Post-radiotherapy cardiotoxicity presents significant clinical challenges, as it negatively affects both overall survival and quality of life. A study of patients with classic Hodgkin’s lymphoma indicated that cardiovascular disease has surpassed the primary cancer and other malignancies as the leading cause of mortality.^7^ In breast cancer, relative risks for specific cardiac conditions and cardiac mortality range from 1.29 to 1.97 across several investigations.^8–10^ In lung cancer, increased radiation dose is associated with reduced overall survival, with a hazard ratio of 1.38 compared with standard-dose treatment.^11^ Darby et al reported a 7.4% increase in the rates of major coronary events per 1 Gy increase in mean heart dose.^12^ However, mean heart dose fails to account for heterogeneous dose distribution within the heart and may not accurately reflect the actual dose delivered to cardiac substructures.^13^ Emerging evidence suggests that dosimetric parameters of specific cardiac substructures better predict cardiotoxicity than whole-heart (WH) dose metrics.^14–22^ Cardiac substructures such as the left ventricle,^16,18^ left atrium,^15^ left anterior descending coronary artery,^14,17,20,21^ and heart valves^19^ have been implicated in radiation-induced cardiac damage. Efforts to spare these critical substructures during treatment planning may reduce the risk of cardiotoxicity while maintaining high dose to targets for tumor control.

The incidence of cardiac complications in patients with lung cancer can reach up to 33%.^23^ Various studies have proposed different dose constraints to mitigate cardiac risk.^24^ Although factors such as pre-existing heart conditions and smoking status also influence risk, an individualized approach for predicting cardiotoxicity is crucial for personalized treatment planning. Because radiation-induced cardiac damage can develop gradually, several biomarkers have emerged as potential indicators of subclinical cardiotoxicity during chemoradiotherapy (CRT).^25^ Cardiac troponin T, released into the serum by damaged cardiac myocytes, serves as a specific marker of myocardial injury in routine cardiology practice.^26^ Elevation in hs-cTnT levels has demonstrated predictive value as an early biomarker of radiation-or chemo-induced myocardial damage.^27–31^ A relationship with clinical cardiotoxicity has, however, not been fully demonstrated yet.

Historically, the labor-intensive process of manually delineating cardiac substructures posed a significant barrier to large-scale dosimetric analyses and cardiotoxicity prediction studies. However, advances in deep-learning–based algorithms have improved the efficiency and consistency of cardiac substructure delineation, producing clinically acceptable contours suitable for dosimetric studies.^32–37^ In this study, we leveraged deep-learning–based auto-segmentation models to facilitate the development of elastic net models for predicting hs-cTnT elevation as a surrogate marker of radiation-induced cardiotoxicity. The models integrated a comprehensive set of inputs, including clinical factors, dose-volume histogram (DVH) parameters, and radiomic and dosiomic features. Dosiomic features are radiomic features extracted from a 3D dose map. We evaluated the predictive performance of WH-based and cardiac substructure-based variables and assessed model robustness through hold-out validation by using data from patients prospectively enrolled in an ongoing clinical trial. This approach holds promise for future integration into personalized treatment planning workflows, enabling more precise cardiotoxicity risk assessment in lung cancer radiotherapy.

## METHODS AND MATERIALS

### Patient cohorts

This study was approved by the institutional review board. The retrospective dataset was obtained from a completed randomized prospective trial comparing intensity-modulated radiation therapy (IMRT) with passive scattering proton therapy (PSPT) in patients with non-small cell lung cancer (NCTxxxxxxxxx).^38^ Of 225 eligible patients, 160 were selected after excluding those who withdrew consent for blood sample donation or lacked sufficient samples for hs-cTnT level analysis. Those patients received either IMRT or PSPT with prescribed doses ranging from 60 Gy to 74 Gy between 2009 and 2014, combined with concurrent chemotherapy. Clinical factors such as sex, age, smoking status, and pre-existing heart diseases were recorded, as detailed in prior publications.^30,38^ Another 74 patients were recruited from an ongoing clinical trial (NCTxxxxxxxxx) starting in 2021, with 57 meeting the same inclusion criteria. Those patients received photon-based treatments, including IMRT or volumetric-modulated arc therapy, or proton-based treatments, including PSPT or intensity-modulated proton therapy (IMPT), with prescribed doses ranging from 60 Gy to 72 Gy. For the majority of patients, tumor internal motion was assessed using four-dimensional computed tomography (4DCT) to generate an internal planning target volume (iPTV), while a minority were treated under breath-hold conditions as per decision of physicians. In IMPT plans, repainting and robust optimization techniques were employed to mitigate range uncertainties and interplay effects associated with respiratory motion. In the current study, the retrospective dataset was used for training and Monte Carlo cross-validation (MCCV) of a prediction model for hs-cTnT elevation, and the prospective dataset served as an independent hold-out validation set for the hs-cTnT elevation prediction model, as illustrated in Figure 1.

**Figure 1.**
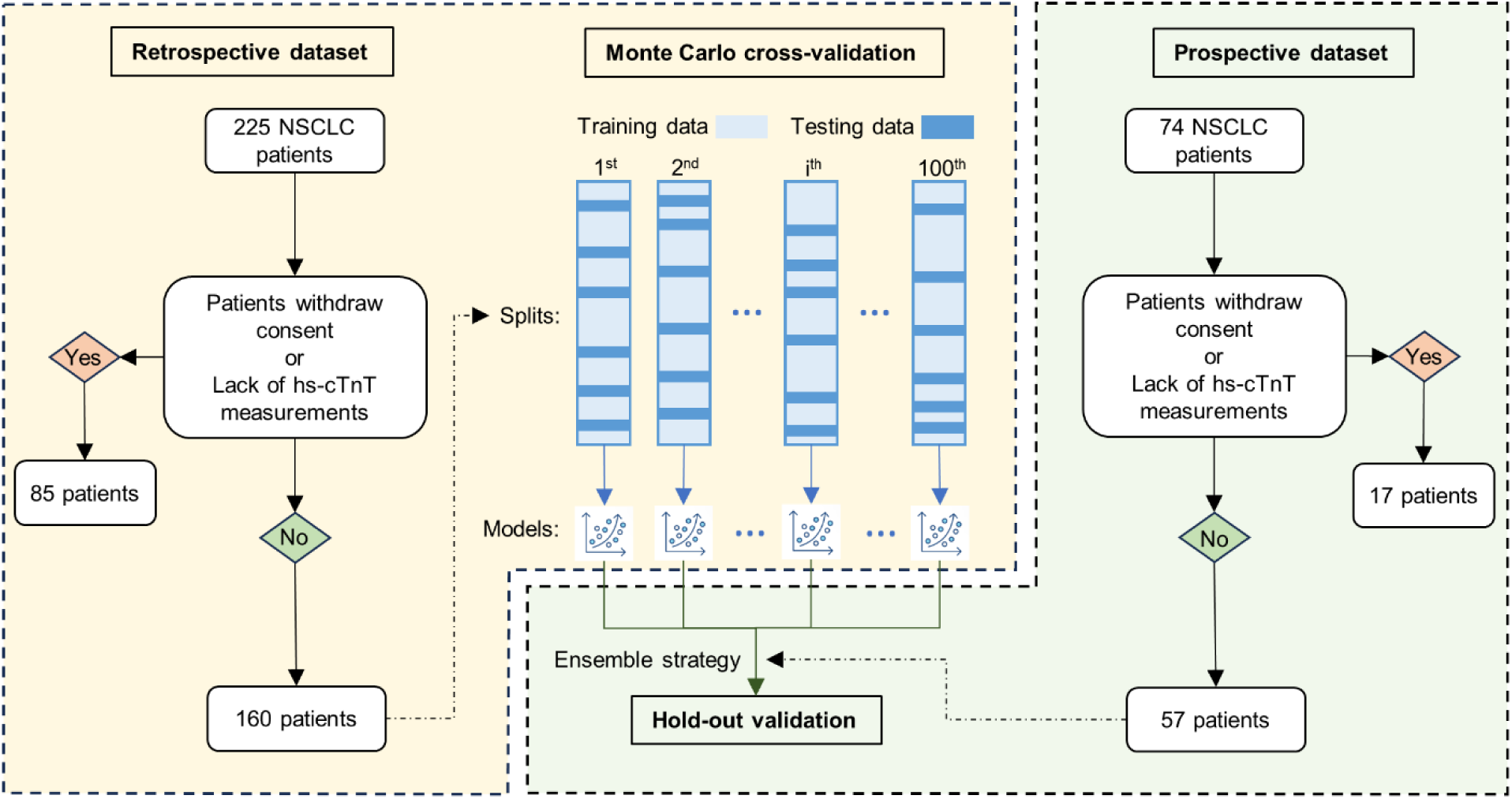
CONSORT diagram with schematic representation of the data collection, Monte Carlo cross-validation, and hold-out validation processes. Abbreviations: NSCLC, non-small cell lung cancer; hs-cTnT, high-sensitivity cardiac troponin T.

**Figure 2.**
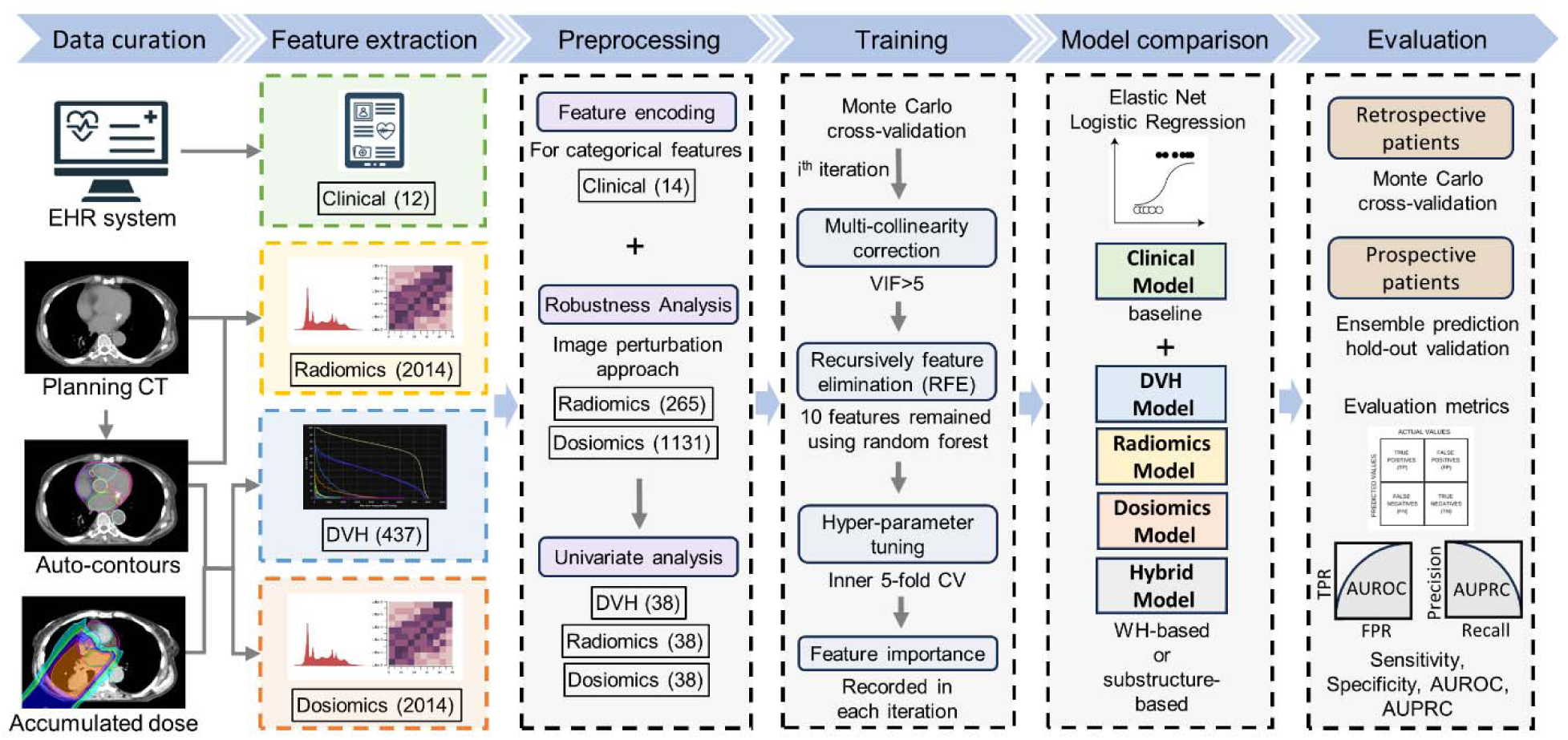
Schematic of study workflow. Abbreviations: EHR, electronic heath record; CT, computed tomography; DVH, dose-volume histogram; VIF, variance inflation factor; CV, cross-validation; WH, whole-heart; AUROC, area under the receiver operating characteristic curve; AUPRC, area under the precision-recall curve; TPR, true-positive rate; FPR, false-positive rate.

### Hs-cTnT measurement

Baseline blood samples were collected within 2 months before CRT for the retrospective dataset and within 2 weeks before CRT for the prospective dataset. Additional samples were obtained 2-3 times during CRT, including during the final week of treatment in both datasets. Serum was separated within 2 hours of collection and stored at −80°C for testing and long-term preservation. The hs-cTnT concentrations in serum were measured in duplicate with an Elecsys TnT Gen 5 STAT assay (Roche Diagnostics, Indianapolis, IN, USA) on a Cobas e411 analyzer. Samples within each dataset were analyzed using the same assay. A delta in hs-cTnT level exceeding 5 ng/L has been linked with a significantly increased risk of cardiac adverse events.^30^ Therefore, in this study, hs-cTnT elevation was defined as a maximum change in hs-cTnT levels exceeding 5 ng/L during CRT and was considered the primary endpoint. Although dichotomizing the hs-cTnT outcome into elevated vs. non-elevated may result in the loss of detailed quantitative information, it mitigates the influence of outlier values and reduces bias introduced by extreme hs-cTnT level changes observed in individual patients.

### Auto-segmentation

An in-house nnU-Net based auto-segmentation model was applied to the planning average 4-dimensional computed tomography images to delineate a total of 19 substructures, including the WH, left atrium, right atrium, left ventricle, right ventricle, ascending aorta, descending aorta, pulmonary artery, pulmonary vein, superior vena cava, inferior vena cava, aortic valve, mitral valve, pulmonary valve, tricuspid valve, left anterior descending coronary artery (LAD), left main coronary artery, left circumflex coronary artery, and right coronary artery.^32^ The model was trained using contours developed according to consensus guidelines informed by the University of Michigan cardiac atlas^39^ and refined with input from our institutional cardiology experts, as detailed in the appendix of the original publication. This model has been extensively evaluated and has achieved an overall rate of clinically acceptable contours of 94%.

### DVH parameters

DVH parameters were extracted from clinically delivered radiotherapy treatment plans by using the open-source Python package “PlatiPy”. For patients with multiple treatment plans or significant plan adaptations, accumulated dose maps were generated by registering the re-simulated CT images to the original planning CT images by deformable image registration, conducted in Raystation 12A (RaySearch Laboratories AB, Stockholm, Sweden). A total of 23 DVH metrics were extracted for each substructure, including minimum dose, mean dose, maximum dose, standard deviation, relative volumes receiving at least x Gy or x Gy(Relative Biologic effectiveness, RBE) (VxGy, including V5Gy and V10Gy to V70Gy in 10-Gy increments), doses received by at least x% of the volume (Dx%, including D5% and D10% to D70% in 10% increments), and dose received by absolute x cc volumes (Dxcc, including D0.1cc, D1.0cc and D5.0cc). In total, 437 DVH parameters were extracted for each patient.

### Radiomic and dosiomic features

A total of 106 radiomic features were extracted from the planning CT for each cardiac substructure. The same feature categories were extracted from the 3D dose maps and denoted as dosiomic features. Feature extraction was done with the open-source Python package “pyradiomics”.^40^ This process resulted in a total of 2,014 radiomic features and 2,014 dosiomic features per patient.

To enhance reliability and generalizability, robustness analysis was done with the retrospective dataset. Unstable features were excluded through an image perturbation approach.^41^ A perturbation chain was applied to each patient to acquire a set of perturbed data including CT images, substructure masks, and dose maps: (1) applying random rotations to CT images, substructure masks, and dose maps to mimic variations in patient positioning; (2) adding random Gaussian noise to CT images to account for differences in imaging systems; (3) performing random volume shrinkage/growth on substructure masks to reflect volume variations in manual delineation; (4) introducing random deformation of masks to represent contour uncertainties. The same features described above were extracted from perturbed CT images, masks, and dose maps. Features with Spearman correlation coefficients of >0.9 from before to after the perturbation were considered robust and were retained for further processing. Detailed procedures for feature extraction and robustness analysis are provided in Appendix A and Appendix B.

### Univariate analysis and pre-selection of variables

Clinical factors, DVH parameters, radiomic features, and dosiomic features were compared between patients with and without hs-cTnT elevation during CRT. The Wilcoxon rank-sum test and Pearson Chi-square test were used for continuous and categorical features, respectively.

Given the large number of DVH, radiomic, and dosiomic features relative to the patient sample size, univariate statistical testing was conducted to identify the most meaningful variables for model input. For each substructure, the two most significant variables within each feature group (DVH, radiomics, dosiomics) were selected based on the lowest *P* values from the Wilcoxon rank-sum tests within the retrospective dataset. These selected features were used to construct elastic net models in the MCCV of the retrospective dataset. A significance level of 0.00015 was applied considering Bonferroni correction for multiple comparisons.

All clinical factors were included in the model construction process, regardless of their individual correlation with hs-cTnT elevation, to account for potential intricate interactions with other features. Statistical test results for the prospective dataset are presented to provide additional insight into the dataset characteristics but were not incorporated into the pre-selection process to prevent data leakage. Before being input into the model, categorical variables among clinical factors were encoded by using a OneHot encoder with the open-source package “scikit-learn”.

### Model construction

We applied a penalized logistic regression, elastic net logistic regression in this study. A Monte Carlo cross validation (MCCV) approach was used to randomly divide the patients from the retrospective dataset into training and testing subsets with a split of 75%/25% for 100 times.

Input variables were grouped as clinical factors, selected DVH parameters, selected robust radiomic features, and selected robust dosimetric features.

To assess the predictive roles of cardiac substructures and variable groups in hs-cTnT elevation, multiple models were constructed for each training-testing split: (1) clinical model: included only clinical factors as a baseline; (2) WH DVH model: clinical factors combined with WH-based DVH parameters; (3) SUB DVH model: clinical factors combined with substructure-based DVH parameters (including the WH); (4) WH radiomic model: clinical factors combined with WH-based radiomic features; (5) SUB radiomic model: clinical factors combined with substructure-based radiomic features; (6) WH dosiomic model: clinical factors combined with WH-based dosiomic features; (7) SUB dosiomic model: clinical factors combined with substructure-based dosiomic features; (8) WH hybrid model: hybrid inputs of clinical factors and all WH-based variables; and (9) SUB hybrid model: hybrid inputs of clinical factors and all substructure-based variables.

### Model training and evaluation

In each iteration, variables within a specific model from the training samples were normalized to a range of 0 to 1 by using a MinMaxScaler. Multicollinearity correction was performed by eliminating features with high collinearity that could negatively influence model performance.

The variance inflation factor (VIF) was used as the elimination criterion, calculated as:

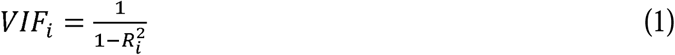

where R*_i_*^2^ represents the unadjusted coefficient of determination for regressing the i-th variable on the remaining variables. Variables with the highest VIFs were iteratively removed until all VIFs were below 5, corresponding to an R^2^ less than 0.8, indicating acceptable collinearity levels. Next, recursive feature elimination (RFE) was applied to select the top 10 variables for model input, balancing the number of inputs with the number of training samples. A default random forest classifier was used within the RFE process to leverage Gini importance and reduce overfitting risks in subsequent logistic regression models.

A unique elastic net logistic regression model was constructed for each of the 100 iterations by using the selected 10 features. An inner 5-fold cross-validation was used to optimize hyperparameters, including the regularization strength, L1/L2 penalty ratio, and solver type. Balanced accuracy was used as the validation metric because of data imbalance in the dataset. The trained model was then applied to the testing samples by using the corresponding selected features, scaled with the same MinMaxScaler. Evaluation metrics, including sensitivity, specificity, area under the receiver operating characteristic curve (AUROC), and area under the precision-recall curve (AUPRC), were calculated based on predicted and ground-truth labels for each iteration. These metrics were aggregated across all iterations for statistical analysis. Model training, testing, data splitting, and hyperparameter tuning were implemented with the open-source package “scikit-learn”.

The MCCV approach increased dataset partitioning, thus reducing estimation variability and enabling direct comparison of performance across different models. Because identical training and testing samples were used across iterations, the Wilcoxon signed-rank tests were applied, which allowed pairwise comparisons of performance metrics between models.

### Feature importance

To evaluate the predictive significance of input variables and identify superior predictors of hs-cTnT elevation, permutation-based feature importance was calculated during model training in each iteration.^42^ Each input variable was randomly permuted 30 times, and the corresponding changes in the training balanced accuracy score were measured as importance. A substantial decrease in the training score after permutation of a specific variable indicated its importance in prediction. Permutation importance values for each variable were recorded across all 100 iterations for further analysis. Variables excluded during multicollinearity correction or RFE were assigned an importance score of zero. For the identified important variable, several common approaches were applied to determine optimal cut-off value regarding hs-cTnT elevation, including Youden index, Euclidean distance, Index of Union, Maximum product of sensitivity and specificity, and minimum P-value.^43^

### Hold-out validation

The prospective patient dataset was used as an independent hold-out set to evaluate the performance of the different models. This dataset was exclusively applied during hold-out validation, ensuring no data leakage and minimizing potential bias. To enhance prediction robustness, an ensemble prediction approach was implemented for each model type by using the 100 elastic net logistic regression models trained during the MCCV. The final probability of hs-cTnT elevation was determined by averaging the logits from each model, calculated as:

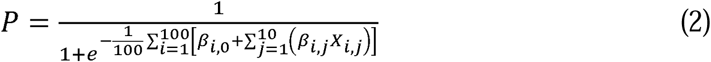

Where X_i,j_ represents the j-th input variable in the i-th model, β_i,j_ represents the j-th coefficient in the i-th model, and β_i,0_ represents the intercept in the i-th model. A probability threshold of 0.5 was used to derive the final prediction. Similarly, sensitivity, specificity, AUROC, and AUPRC were calculated based on the ensembled predictions and ground-truth labels.

## RESULTS

### Patient characteristics: Retrospective vs. prospective

Characteristics of the retrospective and prospective cohorts are summarized in Table 1. Chemotherapy details are omitted, because all patients received the same regimen. No statistically significant differences were observed between the cohorts in terms of sex, tumor stage, tumor histology, tumor location, pre-existing heart disease, Karnofsky Performance Status (KPS) score, treatment modality, or hs-cTnT elevation (the primary endpoint). Notably, compared with the retrospective cohort, prospective patients were older (67.8 yrs vs. 64.7 yrs, *P* = 0.037), had fewer smokers (active: 12% vs. 21%; former: 65% vs. 70%; never: 23% vs. 9%; *P* = 0.013), had higher baseline hs-cTnT levels (11.2 ng/L vs. 7.9 ng/L, *P* = 0.0002), and had lower prescribed radiation doses (66.7 Gy vs. 70.5 Gy, *P* < 0.0001).

**Table 1.** Clinical characteristics in the retrospective and prospective datasets.

|  | Value or No. of Patients (%) |  | *P* Value |
| --- | --- | --- | --- |
|  | Retrospective (n=160) | Prospective (n=57) |  |
| **Age, years, mean (SD)** | 64.7 (8.9) | 67.8 (10.1) | 0.037 |
| **Sex** |  |  | 0.302 |
| **Female** | 78 (49) | 33 (58) |  |
| **Male** | 82 (51) | 24 (42) |  |
| **Race** |  |  | 0.280 |
| **White** | 145 (91) | 48 (84) |  |
| **Other** | 15 (9) | 9 (16) |  |
| **Tumor stage** |  |  | 0.070 |
| **≤IIIA** | 74 (46) | 22 (39) |  |
| **≥IIIB** | 86 (54) | 35 (61) |  |
| **Tumor histology** |  |  | 0.927 |
| **Adeno** | 91 (57) | 34 (60) |  |
| **Squamous** | 52 (32) | 17 (30) |  |
| **Other** | 17 (11) | 6 (10) |  |
| **Tumor location** |  |  | 0.969 |
| **Left/Mediastinal** | 65 (41) | 24 (42) |  |
| **Right** | 95 (59) | 33 (58) |  |
| **Smoking status** |  |  | 0.013 |
| **Never** | 14 (9) | 13 (23) |  |
| **Former** | 112 (70) | 37 (65) |  |
| **Active** | 34 (21) | 7 (12) |  |
| **Pre-existing heart disease** |  |  | 0.456 |
| **No** | 130 (81) | 43 (75) |  |
| **Yes** | 30 (19) | 14 (25) |  |
| **KPS** |  |  | 1.000 |
| **□ 80** | 15 (9) | 6 (10) |  |
| **≥80** | 145 (91) | 51 (90) |  |
| **hs-cTnT baseline, ng/L, mean (SD)** | 7.9 (9.6) | 11.2 (9.5) | <0.0001 |
| **Prescribed dose, Gy, mean (SD)** | 70.5 (4.6) | 66.7 (3.2) | <0.0001 |
| **Modality** |  |  | 0.454 |
| **Photon** | 98 (61) | 31 (54) |  |
| **Proton** | 62 (39) | 26 (46) |  |
| **hs-cTnT elevation** |  |  | 0.904 |
| **No** | 109 (68) | 40 (70) |  |
| **Yes** | 51 (32) | 17 (30) |  |

Continuous variables are reported as mean (standard deviation) and categorical variables are reported as frequency (percentage). *P* values are from Wilcoxon rank sum test for continuous values and Pearson's Chi-square test for categorical variables.
Abbreviations: KPS, Karnofsky Performance Status score; hs-cTnT, high-sensitivity cardiac troponin T.

### Patient characteristics: Photon vs. proton

The incidence of hs-cTnT elevation was non-significantly lower in proton therapy patients in the retrospective cohort (37% vs. 24%, *P* = 0.138) and comparable in the prospective cohort (29% vs. 31%, *P* = 1.000), compared with photon therapy patients. Among the 160 retrospective patients, no significant differences in baseline characteristics were observed between the two treatment groups, except for smoking status (active: 14% vs. 32%; former: 76% vs. 61%; never: 10% vs. 6%; *P* = 0.024). In the prospective dataset, proton therapy patients were significantly older (72.5 yrs vs. 63.9 yrs, *P* = 0.001) and had a higher prevalence of adenocarcinoma histology (adenocarcinoma: 73% vs. 49%; squamous cell carcinoma: 27% vs. 32%; other: 0% vs. 6%; *P* = 0.037). Detailed comparisons of patient characteristics are provided in Appendix C and Appendix D. Given the relatively minor differences in hs-cTnT elevation rates and patient characteristics, both photon and proton therapy patients were included in the same predictive model, with treatment modality incorporated as an input variable to assess its potential impact on cardiotoxicity prediction.

### Robustness analysis and pre-selection of variables

After the perturbation chain analysis, 265 of 2,014 radiomic features and 1,131 of 2014 dosiomic features were identified as being robust against image perturbations. Detailed robust feature counts per substructure are provided in Appendix B. During variable pre-selection, two DVH parameters, two radiomic features, and two dosiomic features were chosen for each substructure based on the statistical distribution observed in the retrospective dataset. A comparative summary of pre-selected WH-based and LAD-based variables is presented in Table 2. The pre-selection process was conducted exclusively on the retrospective dataset; however, to provide additional context on population differences, prospective dataset statistics are also included in Table 2. In the retrospective dataset, all WH-based variables demonstrated no significant association with hs-cTnT elevation (*P* > 0.001, significance level 0.00015), indicating their limited predictive value. In comparison, retrospective patients with hs-cTnT elevation (n=51) had significantly higher LAD D1.0cc (31.2 Gy vs. 13.8 Gy, *P* < 0.0001) and LAD V20Gy (38.7% vs. 13.2%, *P* < 0.0001) compared with those without elevation (n=109). The dosiomic feature LAD glszm_ZoneEntropy (which quantifies the complexity of zone sizes with different dose levels) also significantly distinguished the two patient groups. Notably, the selected dosiomic features LAD glszm_ZoneEntropy and LAD firstorder_90Percentile were strongly correlated with LAD V20Gy, with Spearman correlation coefficients of 0.84 and 0.91. A comparison between the retrospective and prospective cohorts revealed that prospective patients received significantly lower WH V20Gy (10.7% vs. 21.7%, *P* < 0.0001), although no significant differences in LAD-based variables were observed (*P*>0.001). However, both WH-and LAD-based variables showed no significant differences between prospective patients with and without hs-cTnT elevation, highlighting substantial variability within the retrospective and prospective datasets. Comprehensive tables of pre-selected DVH, radiomic, and dosiomic features for each cardiac substructure are presented in Appendices F-H.

**Table 2.**
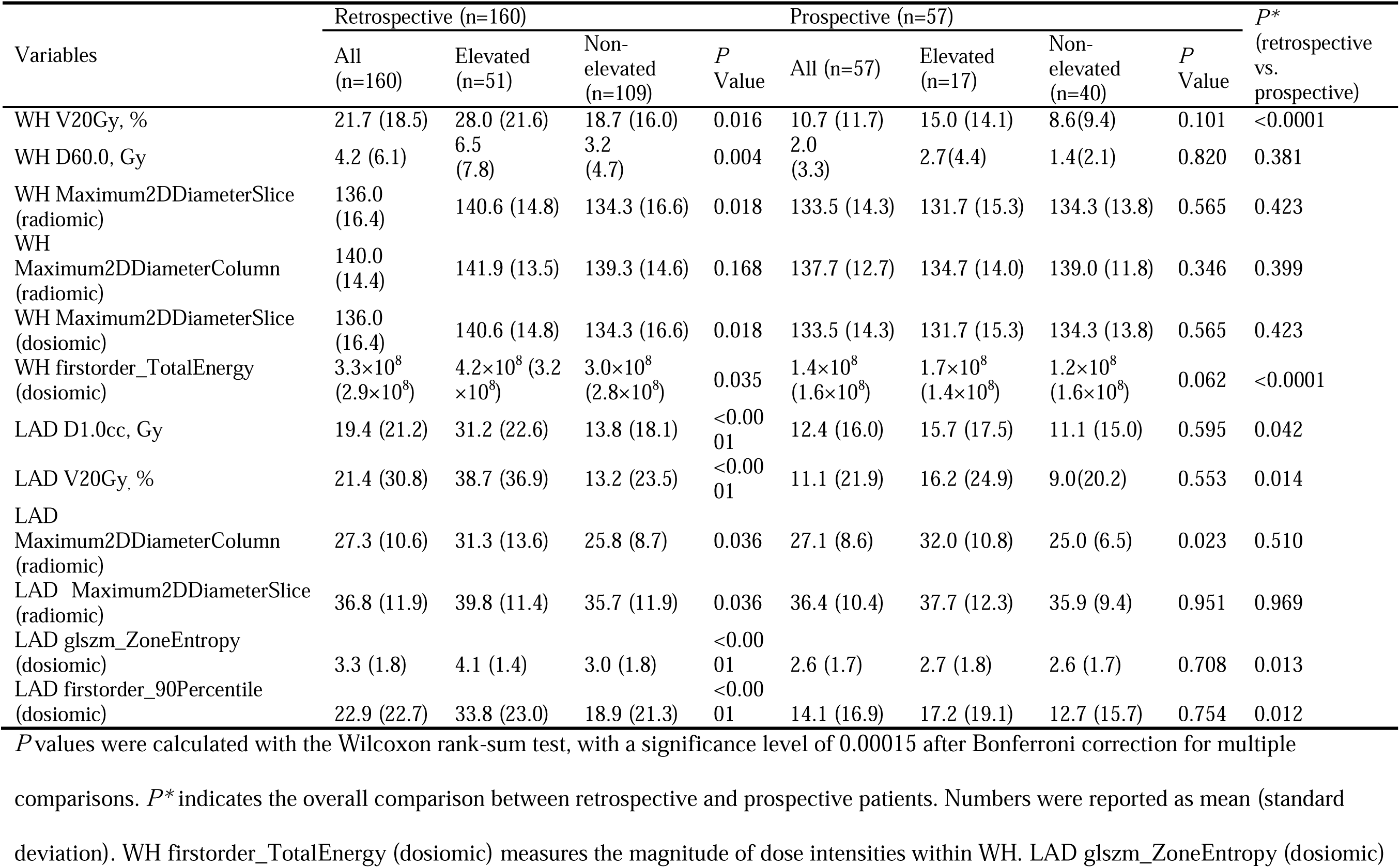
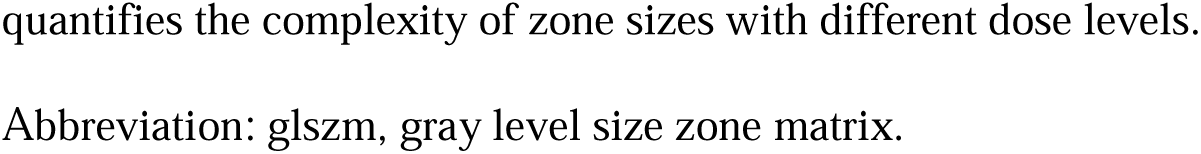
Comparison of whole heart (WH)-based and left anterior descending coronary artery (LAD)-based variables between patients with and without hs-cTnT elevation in the retrospective and prospective cohorts.

| Variables | Retrospective (n=160) |  |  |  | Prospective (n=57) |  |  |  | *P* *<br>(retrospective<br>vs.<br>prospective) |
| --- | --- | --- | --- | --- | --- | --- | --- | --- | --- |
|  | All<br>(n=160) | Elevated<br>(n=51) | Non-elevated<br>(n=109) | *P*<br>Value | All (n=57) | Elevated<br>(n=17) | Non-elevated<br>(n=40) | *P*<br>Value |  |
| WH V20Gy, % | 21.7 (18.5) | 28.0 (21.6) | 18.7 (16.0) | 0.016 | 10.7 (11.7) | 15.0 (14.1) | 8.6(9.4) | 0.101 | <0.0001 |
| WH D60.0, Gy | 4.2 (6.1) | 6.5<br>(7.8) | 3.2<br>(4.7) | 0.004 | 2.0<br>(3.3) | 2.7(4.4) | 1.4(2.1) | 0.820 | 0.381 |
| WH Maximum2DDiameterSlice<br>(radiomic) | 136.0<br>(16.4) | 140.6 (14.8) | 134.3 (16.6) | 0.018 | 133.5 (14.3) | 131.7 (15.3) | 134.3 (13.8) | 0.565 | 0.423 |
| WH<br>Maximum2DDiameterColumn<br>(radiomic) | 140.0<br>(14.4) | 141.9 (13.5) | 139.3 (14.6) | 0.168 | 137.7 (12.7) | 134.7 (14.0) | 139.0 (11.8) | 0.346 | 0.399 |
| WH Maximum2DDiameterSlice<br>(dosiomic) | 136.0<br>(16.4) | 140.6 (14.8) | 134.3 (16.6) | 0.018 | 133.5 (14.3) | 131.7 (15.3) | 134.3 (13.8) | 0.565 | 0.423 |
| WH firstorder_TotalEnergy<br>(dosiomic) | 3.3×10<sup>8</sup><br>(2.9×10<sup>8</sup>) | 4.2×10<sup>8</sup> (3.2<br>×10<sup>8</sup>) | 3.0×10<sup>8</sup><br>(2.8×10<sup>8</sup>) | 0.035 | 1.4×10<sup>8</sup><br>(1.6×10<sup>8</sup>) | 1.7×10<sup>8</sup><br>(1.4×10<sup>8</sup>) | 1.2×10<sup>8</sup><br>(1.6×10<sup>8</sup>) | 0.062 | <0.0001 |
| LAD D1.0cc, Gy | 19.4 (21.2) | 31.2 (22.6) | 13.8 (18.1) | <0.00<br>01 | 12.4 (16.0) | 15.7 (17.5) | 11.1 (15.0) | 0.595 | 0.042 |
| LAD V20Gy, % | 21.4 (30.8) | 38.7 (36.9) | 13.2 (23.5) | <0.00<br>01 | 11.1 (21.9) | 16.2 (24.9) | 9.0(20.2) | 0.553 | 0.014 |
| LAD<br>Maximum2DDiameterColumn<br>(radiomic) | 27.3 (10.6) | 31.3 (13.6) | 25.8 (8.7) | 0.036 | 27.1 (8.6) | 32.0 (10.8) | 25.0 (6.5) | 0.023 | 0.510 |
| LAD Maximum2DDiameterSlice<br>(radiomic) | 36.8 (11.9) | 39.8 (11.4) | 35.7 (11.9) | 0.036 | 36.4 (10.4) | 37.7 (12.3) | 35.9 (9.4) | 0.951 | 0.969 |
| LAD glszm_ZoneEntropy<br>(dosiomic) | 3.3 (1.8) | 4.1 (1.4) | 3.0 (1.8) | <0.00<br>01 | 2.6 (1.7) | 2.7 (1.8) | 2.6 (1.7) | 0.708 | 0.013 |
| LAD firstorder_90Percentile<br>(dosiomic) | 22.9 (22.7) | 33.8 (23.0) | 18.9 (21.3) | <0.00<br>01 | 14.1 (16.9) | 17.2 (19.1) | 12.7 (15.7) | 0.754 | 0.012 |

*P* values were calculated with the Wilcoxon rank-sum test, with a significance level of 0.00015 after Bonferroni correction for multiple
comparisons. *P*\* indicates the overall comparison between retrospective and prospective patients. Numbers were reported as mean (standard
deviation). WH firstorder\_TotalEnergy (dosiomic) measures the magnitude of dose intensities within WH. LAD glszm\_ZoneEntropy (dosiomic)
quantifies the complexity of zone sizes with different dose levels.
Abbreviation: glszm, gray level size zone matrix.

It is noteworthy that factors commonly considered as cardiotoxicity risk indicators, such as baseline hs-cTnT levels (8.1 vs. 7.8 ng/L, *P* = 0.095) and the presence of pre-existing heart disease (19.6% vs. 18.3%, *P* = 1.000), did not show statistically significant differences between patients with and without hs-cTnT elevation. Nonetheless, all clinical variables were retained as model inputs to account for potential complex interaction effects among predictors.

### Model evaluation

The sensitivity, specificity, AUROC, and AUPRC results from MCCV and hold-out validation are summarized in Table 3. The baseline clinical model achieved a mean AUROC of 0.66 in MCCV. Incorporating WH-based variables marginally improved the predictive performance of the model (AUROC: WH DVH, 0.67, *P* = 0.0003; WH radiomics, 0.67, *P* = 0.030; WH dosiomics, 0.66, *P* = 0.083). In contrast, incorporating substructure-based DVH parameters and dosiomic features significantly improved the model’s performance, yielding mean AUROCs of 0.71 (*P* < 0.0001) for clinical plus substructure DVH parameters and 0.70 (*P* < 0.0001) for clinical plus substructure dosiomic features. However, adding substructure-based radiomic features reduced the AUROC to 0.57 (*P* < 0.0001). The hybrid model incorporating substructure-based features (SUB hybrid) performed worse than the SUB DVH model (AUROC: 0.68 vs. 0.71, *P* < 0.0001). In comparison, the WH hybrid model, combining WH-based DVH parameters, radiomic, and dosiomic features, achieved an AUROC of 0.70, comparable to the SUB DVH model (*P* = 0.051). Nevertheless, in hold-out validation, performance of the clinical baseline model and all WH-based models was limited (AUROC: clinical, 0.45; WH DVH, 0.50; WH radiomics, 0.48; WH dosiomics, 0.48; WH hybrid, 0.51), indicating poor generalizability across datasets. Conversely, the SUB DVH model achieved an AUROC of 0.60, with a sensitivity of 0.47 and specificity of 0.75. The SUB dosiomics model performed worse (AUROC: 0.56), despite high correlations between selected dosiomic features and DVH parameters in the retrospective dataset. The SUB radiomics model achieved an AUROC of 0.61; however, its lower MCCV performance suggests that its higher AUROC in hold-out validation resulted from dataset variability rather than true predictive strength. These trends also applied to AUPRC, which is considered more robust for imbalanced datasets.

**Table 3.**
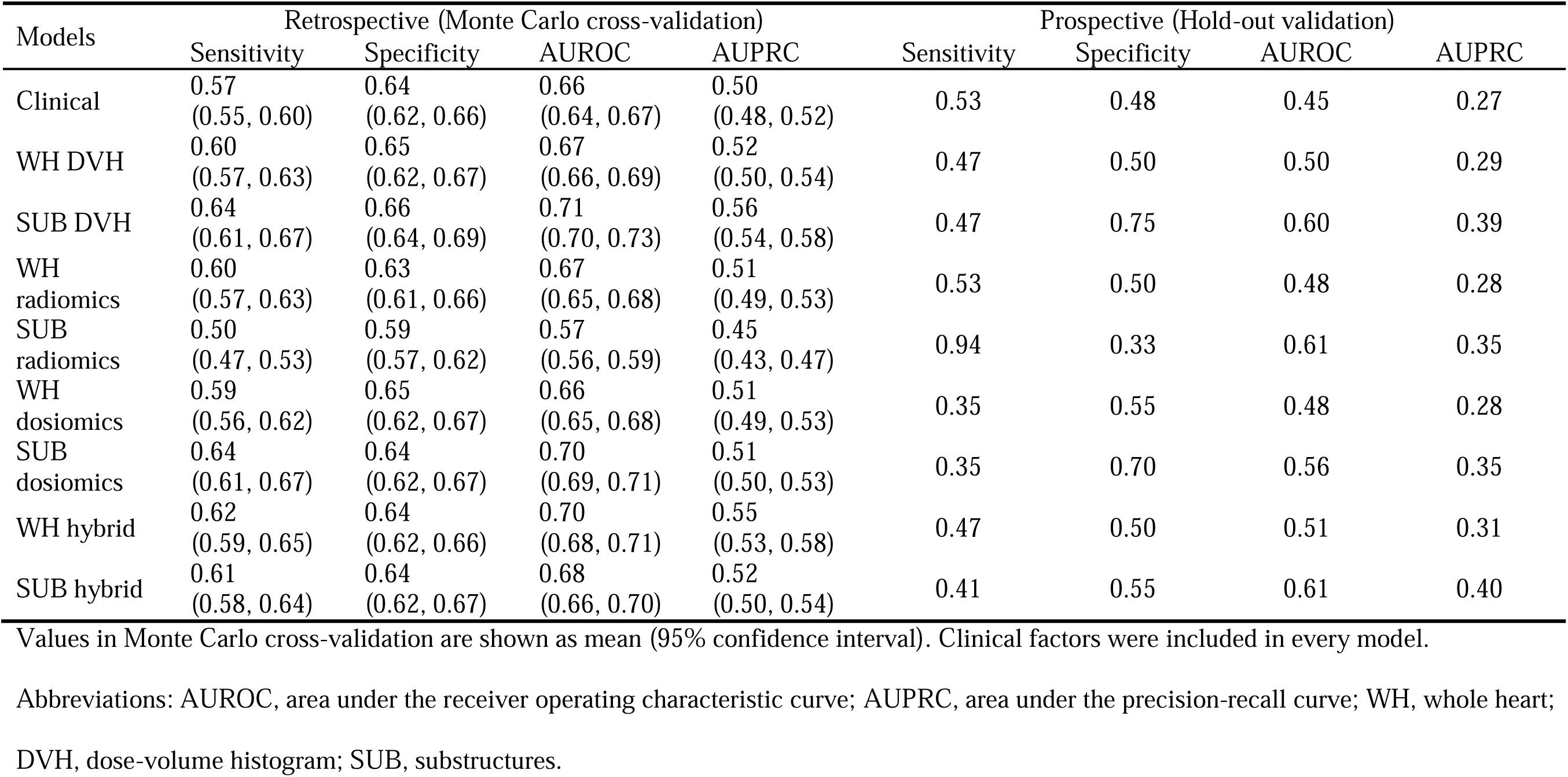
Comparison of model performance in Monte Carlo cross-validation (within the retrospective cohort) and in hold-out validation (within the prospective cohort)

| Models | Retrospective (Monte Carlo cross-validation) |  |  |  | Prospective (Hold-out validation) |  |  |  |
| --- | --- | --- | --- | --- | --- | --- | --- | --- |
|  | Sensitivity | Specificity | AUROC | AUPRC | Sensitivity | Specificity | AUROC | AUPRC |
| Clinical | 0.57<br>(0.55, 0.60) | 0.64<br>(0.62, 0.66) | 0.66<br>(0.64, 0.67) | 0.50<br>(0.48, 0.52) | 0.53 | 0.48 | 0.45 | 0.27 |
| WH DVH | 0.60<br>(0.57, 0.63) | 0.65<br>(0.62, 0.67) | 0.67<br>(0.66, 0.69) | 0.52<br>(0.50, 0.54) | 0.47 | 0.50 | 0.50 | 0.29 |
| SUB DVH | 0.64<br>(0.61, 0.67) | 0.66<br>(0.64, 0.69) | 0.71<br>(0.70, 0.73) | 0.56<br>(0.54, 0.58) | 0.47 | 0.75 | 0.60 | 0.39 |
| WH radiomics | 0.60<br>(0.57, 0.63) | 0.63<br>(0.61, 0.66) | 0.67<br>(0.65, 0.68) | 0.51<br>(0.49, 0.53) | 0.53 | 0.50 | 0.48 | 0.28 |
| SUB radiomics | 0.50<br>(0.47, 0.53) | 0.59<br>(0.57, 0.62) | 0.57<br>(0.56, 0.59) | 0.45<br>(0.43, 0.47) | 0.94 | 0.33 | 0.61 | 0.35 |
| WH dosiomics | 0.59<br>(0.56, 0.62) | 0.65<br>(0.62, 0.67) | 0.66<br>(0.65, 0.68) | 0.51<br>(0.49, 0.53) | 0.35 | 0.55 | 0.48 | 0.28 |
| SUB dosiomics | 0.64<br>(0.61, 0.67) | 0.64<br>(0.62, 0.67) | 0.70<br>(0.69, 0.71) | 0.51<br>(0.50, 0.53) | 0.35 | 0.70 | 0.56 | 0.35 |
| WH hybrid | 0.62<br>(0.59, 0.65) | 0.64<br>(0.62, 0.66) | 0.70<br>(0.68, 0.71) | 0.55<br>(0.53, 0.58) | 0.47 | 0.50 | 0.51 | 0.31 |
| SUB hybrid | 0.61<br>(0.58, 0.64) | 0.64<br>(0.62, 0.67) | 0.68<br>(0.66, 0.70) | 0.52<br>(0.50, 0.54) | 0.41 | 0.55 | 0.61 | 0.40 |

Values in Monte Carlo cross-validation are shown as mean (95% confidence interval). Clinical factors were included in every model.
Abbreviations: AUROC, area under the receiver operating characteristic curve; AUPRC, area under the precision-recall curve; WH, whole heart;
DVH, dose-volume histogram; SUB, substructures.

### Feature importance

The permutation importance of input variables across different models in MCCV is summarized in Figure 3. In the clinical model, tumor location emerged as the most influential predictor for predicting hs-cTnT elevation. Similarly, tumor location remained the dominant feature across all WH-based models. WH V20Gy and WH D60.0 exhibited median importance values of zero in the WH DVH model, indicating limited predictive relevance. In the SUB DVH model, LAD V20Gy was identified as the most critical predictor. Similarly, the SUB dosiomics model highlighted LAD glszm_ZoneEntropy as a key variable, with both features ranking among the top predictors in the SUB hybrid model. Given the superior performance of the SUB DVH model in both MCCV and hold-out validation, LAD V20Gy was considered the most dominant predictor among all input variables. Optimal cut-off LAD V20Gy for hs-cTnT elevation was calculated among retrospective patients as 0.3%, 1.9%, 5.0%, 1.1%, and 0.2%, using Youden index, Euclidean distance, Index of Union, maximum product of sensitivity and specificity, and minimum P-value approaches, respectively.

**Figure 3.**
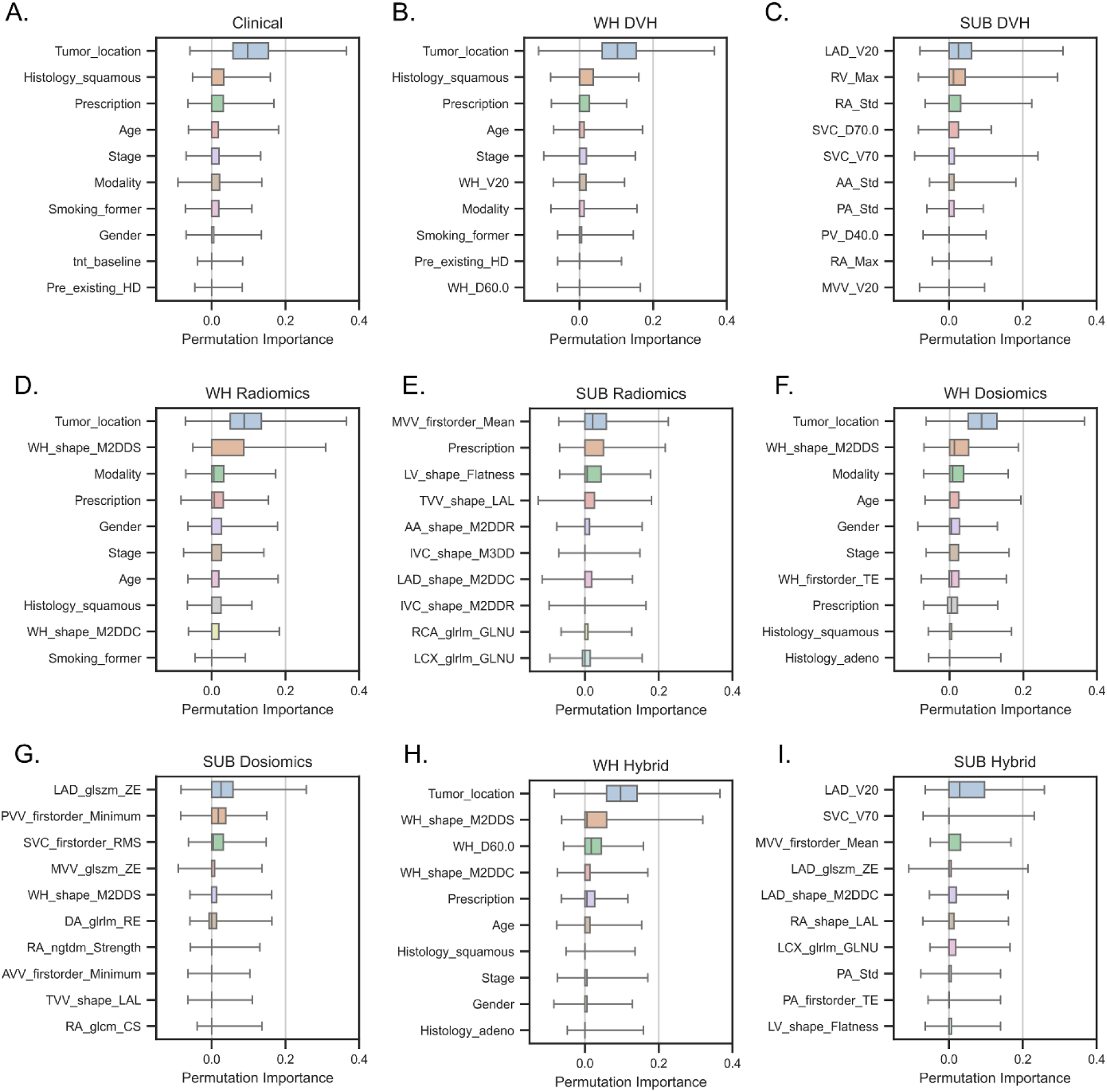
Importance of input variables in permutations in the (A) clinical model, (B) whole-heart dose-volume histogram (WH DVH) model, (C) substructure (SUB) DVH model, (D) WH radiomics model, (E) SUB radiomics model, (F) WH dosiomics model, (G) SUB dosiomics model, (H) WH hybrid model, and (I) SUB hybrid model. Error bars indicate the variability in feature importance across folds of Monte Carlo cross-validation. “Prescription” refers to the prescribed dose to the target volume, and “Modality” indicates the treatment type (photon or proton). Abbreviations: pre_existing HD, pre-existing heart disease; LAD, left anterior descending coronary artery; LV, left ventricle; RV, right ventricle; RA, right atrium; SVC, superior vena cava; IVC, inferior vena cava; AA, ascending aorta; DA, descending aorta; PA, pulmonary artery; PV, pulmonary vein; PVV, pulmonary valve; MVV, mitral valve; TVV, tricuspid valve; AVV, aortic valve; RCA, right coronary artery; LCX, left circumflex coronary artery; Std, standard deviation; glrlm, gray level run length matrix; glszm, gray level size zone matrix; ngtdm, neighboring gray tone difference matrix; glcm, gray level co-occurrence matrix; M2DDS, maximum 2D diameter slice; M2DDC, maximum 2D diameter column; M2DDR, maximum 2D diameter row; M3DD, maximum 3D diameter; GLNU, gray-level non-uniformity; LAL, least axis length; TE, total energy; ZE, zone entropy.

To reduce the risk of multicollinearity and enhance clinical interpretability, predictive models should favor a smaller set of dominant input features. A simplified model constructed using the top five features identified in Figure 3C (LAD V20Gy, RV Max, RA Std, SVC D70, SVC V70) achieved an AUROC of 0.74 in MCCV and 0.60 in hold-out validation.

## DISCUSSION

In this study, we undertook a comprehensive comparison between WH-based and cardiac substructure-based variables for predicting hs-cTnT elevation. We evaluated the predictive performance of DVH parameters, radiomic, and dosiomic features by using both MCCV and hold-out validation. Our findings demonstrate that incorporating substructure-based DVH parameters alongside clinical factors yielded the best performance in MCCV and superior robustness in hold-out validation. Current clinical guidelines predominantly use WH constraints, such as mean heart dose < 26 Gy or Heart V30Gy < 46%, as recommended by the Quantitative Analyses of Normal Tissue Effects in the Clinic (QUANTEC) guidelines.^44^ However, our findings reveal that WH-based dose metrics are poor predictors of hs-cTnT elevation, likely because of their inability to capture localized dose distributions. Notably, dose metrics specific to the LAD were more strongly associated with hs-cTnT elevation than traditional WH dose metrics, such as WH V20Gy. In our feature analysis, tumor location emerged as the most critical factor in all WH-based models, whereas WH-based variables failed to reflect specific treatment sites. Tumor location likely serves as a surrogate for substructure doses, as evidenced by its significant correlations with specific metrics, such as LAD V20Gy (Spearman *r* = 0.58) and SVC V70Gy (*r* = –0.55), and its inferior predictive value. Our results underscore the importance of sparing the LAD during lung cancer treatment planning to minimize the risk of radiation-induced cardiotoxicity.

Identification of heart dose-volume parameters has been inconsistent across different studies,^45^ which could stem from the variability in the ability of the WH DVH parameters to capture substructure-specific doses. Although WH DVH parameters may correlate with substructure doses under fixed tumor locations (e.g., breast radiotherapy), this relationship weakens in more heterogeneous treatment scenarios, such as lung and esophageal cancers. To mitigate this limitation, we examined a comprehensive set of 19 cardiac substructures, including chambers, great vessels, valves, and coronary arteries. This approach reduced the risk of confounding effects and enabled the identification of LAD V20Gy as a robust predictor after correction for multicollinearity. In a dosimetric study of breast cancer patients using a ≥30% increase in hs-cTnT from baseline as the endpoint, LAD V20Gy and V15Gy were significantly higher among patients with elevated hs-cTnT levels (21% incidence).^46^ While that study also identified WH V20Gy as a significant discriminator, our results indicate that WH V20Gy is not a reliable predictor of hs-cTnT elevation in lung cancer patients. This discrepancy may be partly due to the more uniform treatment setup in breast cancer, where WH V20Gy could serve as a surrogate for LAD V20Gy. Our findings further support the importance of LAD sparing, consistent with prior literature.^47^

Proton therapy is generally associated with reduced cardiac dose in lung cancer patients.

In our dosimetric analysis, proton therapy resulted in significantly lower whole-heart doses compared to photon therapy (e.g., WH D60.0: 0.7 Gy vs. 6.5 Gy in the retrospective cohort; 0.3 Gy vs. 3.5 Gy in the prospective cohort; *P* < 0.0001 for both). Among the retrospective patients used for model training, the LAD V20Gy was significantly lower in proton patients than in photon patients (16.6% vs. 34.3%, *P* = 0.0001). In contrast, this difference was not statistically significant in the prospective cohort (9.0% vs. 16.2%, *P* = 0.553). These findings are consistent with the slightly lower incidence (although non-significant) of hs-cTnT elevation observed in proton patients within the retrospective cohort, and the comparable incidence rates between modalities in the prospective cohort. Overall, treatment modality did not emerge as a significant predictor in the model.

Radiomics has had significant roles in studies of outcome prediction in radiotherapy and has demonstrated potential for enhancing predictive performance.^48,49^ Recently, Talebi et al reported that radiomic features significantly enhanced cardiotoxicity prediction in breast cancer based on a cohort of 83 patients.^50^ However, reproducibility and generalizability remain critical challenges owing to the high-dimensional feature space, which increases the risk of data overfitting. In this study, we used an image perturbation strategy to identify robust features resilient to simulated variations in image acquisition and contour delineation. As a result, only 13% of radiomic features and 56% of dosiomic features were retained before feature selection and model construction. Despite an improvement in predictive performance by incorporating radiomic and dosiomic features during MCCV (an increase in AUROC from 0.67 to 0.70), this enhancement did not translate to hold-out validation (AUROC 0.50 vs. 0.51). Moreover, although selected dosiomic features were highly correlated with substructure DVH parameters, the SUB dosiomic model exhibited limited robustness in hold-out validation. These findings underscore the necessity of validating radiomics-based models across independent datasets to ensure their clinical applicability. Variability in imaging protocols, treatment planning quality, and delivery processes may further complicate the reliable application of prediction models in clinical practice.

In this study, we used a clinically validated deep learning-based auto-segmentation model to delineate cardiac substructures from planning CT images. This model demonstrated high geometric and dosiomic accuracy, offering several advantages for dosimetric studies. Notably, it reduced inter-observer variability and improved the consistency of dosimetric metrics. Manual segmentation of cardiac structures, particularly small substructures like the LAD, is prone to substantial variability.^51^ The use of auto-segmentation not only enhances contour consistency but also improves model performance, as evidenced by recent studies demonstrating superior prediction accuracy using auto-contours compared with manual contours.^52^ The adoption of auto-segmentation in both retrospective and prospective patient cohorts also minimized potential deviations caused by the decade-long gap between updates of the contouring guidelines.

Moreover, auto-segmentation mitigated the labor-intensive and time-consuming process of manual contouring, facilitating large-scale retrospective dosimetric analyses. Our results hold promise for the application of auto-segmentation models in advancing future large-scale cardiotoxicity research.

One acknowledged limitation of the current study is that it was conducted at a single institution, which may limit the generalizability of the findings. Expanding the analysis to a multi-institutional dataset would enhance the robustness and external validity of the results. Nonetheless, the inclusion of newly enrolled prospective patients as a hold-out validation cohort improved the reliability of the model evaluation. Another limitation is the relatively small numbers of patients, which was driven by the requirement for hs-cTnT assays and thereby restricting the number of evaluable cases. To address this constraint, we used MCCV to reduce random biases introduced during data splitting and to enable direct pairwise comparisons between models incorporating different variables.

Our results demonstrated moderate cross-validation AUROC and low hold-out validation AUROC, even with the best-performing model, suggesting the presence of unaccounted underlying factors. The elevation of hs-cTnT may not be solely attributable to radiation dose and the clinical variables included here, underscoring the need for broader patient data curation in clinical trials to enhance prediction models. Moreover, hs-cTnT elevation is not always a definitive indicator of underlying cardiac disease.^26^ Accurately assessing adverse cardiac events requires long-term follow-up of prospectively enrolled patients, which is ongoing. We plan to validate our workflow by using actual cardiac adverse events as outcomes in the future. In this study, a fixed RBE of 1.1 was applied for proton dose calculations. This simplification may introduce uncertainties in outcome analysis and potentially underestimate the cardiovascular risk associated with proton therapy. Further investigation into the tissue-specific RBE of protons, particularly in cardiovascular structures, is warranted to determine whether incorporating variable RBE values could improve model performance and cardiotoxicity risk prediction.

Despite these challenges, predicting hs-cTnT elevation enables early intervention to improve treatment plan quality, which ultimately benefits patients. Future research will focus on integrating this predictive model into treatment planning to optimize radiotherapy plans, aiming to reduce cardiotoxicity risk and evaluate the associated clinical benefits.

## CONCLUSIONS

We demonstrated that cardiac substructure DVH parameters provide superior predictive power and robustness compared with WH-based variables for predicting hs-cTnT elevation in radiotherapy for non-small cell lung cancer. LAD V20Gy emerged as a significant predictor of elevated hs-cTnT levels and perhaps of radiation-induced cardiotoxicity. The optimal cut-off value of LAD V20Gy ranged from 0.2% to 5% using different approaches. This model offers valuable insights for cardiotoxicity risk assessment, facilitating the development of personalized treatment plans aimed at minimizing cardiotoxicity in affected patients.

## Data Availability

All data produced in the present study are available upon reasonable request to Zhongxing Liao.

## Notes

### Competing Interest Statement

The authors have declared no competing interest.

### Funding Statement

This work was supported in part by the National Institutes of Health through Research Project Grant R01HL157273-03 and Cancer Center Support (Core) Grant P30016672, start-up funds from MD Anderson Cancer Center, and a grant from the Radiation Oncology Institute (ROI2022-9133).

### Author Declarations

IRB of The University of Texas MD Anderson Cancer Center gave ethical approval for this work.

