## Supplemental Materials for "Radiation doses to cardiac substructures predict elevation in high-sensitivity cardiac troponin T (hs-cTnT) levels in radiotherapy for lung cancer"

**Appendix A: Radiomic and dosiomic feature extraction**

Before feature extraction, the CT images, substructure contours, and 3D dose maps were resampled to an isotropic resolution of 1 mm × 1 mm × 1 mm by using the nearest neighborhood algorithm. The features listed below were extracted from CT images (referred to radiomic features) and dose maps (referred to dosiomic features):

- 18 First-order features: Energy, Total Energy, Entropy, Minimum, Maximum, Mean, Median, Range, Mean Absolute Deviation, Robust Mean Absolute Deviation, Root Mean Square, Variance, Skewness, Kurtosis, Uniformity, 10th Percentile, 90th Percentile, Interquartile Range.
- 13 3D-shape features: Mesh Volume, Voxel Volume, Surface Area, Surface Area to Volume Ratio, Sphericity, Maximum 3D Diameter, Maximum 2D Diameter Slice, Maximum 2D Diameter Column, Maximum 2D Diameter Row, Major Axis Length, Minor Axis Length, Least Axis Length, Flatness.
- 24 gray-level co-occurrence matrix (GLCOM) -based features: Autocorrelation, Cluster Prominence, Cluster Shade, Cluster Tendency, Contrast, Correlation, Difference Average, Difference Entropy, Difference Variance, Joint Average, Joint Energy, Joint Entropy, Inverse Difference (ID), Inverse Difference Normalized (IDN), Inverse Difference Moment (IDM), Inverse Difference Moment Normalized (IDMN), Inverse Variance, Maximum Probability, Sum Average, Sum Entropy, Sum of Squares, Maximal Correlation Coefficient (MCC), Informational Measure of Correlation (IMC) 1, Informational Measure of Correlation (IMC) 2.
- 16 gray-level run length matrix (GLRLM) based features: Short Run Emphasis (SRE), Long Run Emphasis (LRE), Gray-Level Non-Uniformity (GLNU), Gray-Level Non-Uniformity Normalized (GLNUN), Run Length Non-Uniformity (RLNU), Run Length Non-Uniformity Normalized (RLNUN), Run Percentage (RP), Run Entropy, Run Variance, Gray-Level Variance (GLV), Low Gray-Level Run Emphasis (LGRE), High Gray-Level Run Emphasis (HGRE), Short Run Low Gray-Level Emphasis (SRLGLE), Short Run High Gray-Level Emphasis (SRHGLE), Long Run Low Gray-Level Emphasis (LRLGLE), Long Run High Gray-Level Emphasis (LRHGLE).
- 16 gray-level size zone matrix (GLSZM) based features: Small Area Emphasis (SAE), Large Area Emphasis (LAE), Gray-Level Non-Uniformity (GLNU), Gray-Level Non-Uniformity Normalized (GLNUN), Gray-Level Variance (GLV), Size Zone Non-Uniformity (ZSNU), Size Zone Non-Uniformity Normalized (ZSNUN), Zone Percentage, Zone Entropy, Zone Variance, Low Gray-Level Zone Emphasis (LGZE), High Gray-Level Zone Emphasis (HGZE), Small Area Low Gray-Level Emphasis (SALGLE), Small Area High Gray-Level Emphasis (SAHGLE), Large Area Low Gray-Level Emphasis (LALGLE), Large Area High Gray-Level Emphasis (LAZHGLE).
- 14 gray-level dependence matrix (GLDM) based features: Small Dependence Emphasis (SDE), Large Dependence Emphasis (LDE), Gray-Level Non-Uniformity (GLNU), Gray-Level Variance (GLV), Dependence Non-Uniformity (DNU), Dependence Non-Uniformity Normalized (DNUN), Dependence Entropy, Dependence Variance, Low Gray-Level Emphasis (LGLE), High Gray-Level Emphasis (HGLE), Small Dependence Low Gray-Level Emphasis (SDLGLE), Small Dependence High Gray-Level Emphasis (SDHGLE), Large Dependence Low Gray-Level Emphasis (LDLGLE), Large Dependence High Gray-Level Emphasis (LDHGLE).
- 5 neighboring gray-tone difference matrix (NGTDM) based features: Coarseness, Contrast, Busyness, Complexity, Strength.

For the calculation of intensity histograms and other matrices, CT intensities were binned by using a 25 Hounsfield unit bin length, whereas dose maps had a 2.5 Gy bin length.

**Appendix B: Robustness analysis of radiomic and dosiomic features**

In robustness analysis, we applied an image perturbation approach to mimic test-retest procedures. The CT image and dose map from each patient were processed as follows:

1. Rotation: CT images, dose maps, and contours were randomly rotated with an angle ranging from –12 degrees to 12 degrees.
2. Noise: Random gaussian noise was applied to CT images with a standard deviation ranging from 0.1 pixel to 2 pixels.
3. Volume adaptation: The volumes of substructure contours were randomly shrunk or expanded with a volume ratio ranging from 0.8 to 1.2, using "scipy.ndimage.binary_erosion" or "scipy.ndimage.binary_dilation".
4. Contour randomization: The substructure contours were randomly deformed by using an in-house function with B-spline standard deviations ranging from 1 pixel to 4 pixels.

After the perturbation, the same radiomic features were extracted from perturbed CT images and dose maps by using perturbed contours. Spearman correlation factors were calculated between the same features extracted before and after image perturbation. A Spearman correlation higher than 0.9 indicates robust features against perturbation uncertainties. In total, 265 of 2014 radiomic features and 1131 of dosiomic features were considered robust (Figure A1). For whole-heart (WH), only 9 radiomic features and 44 dosiomic features were robust. By comparison, more features extracted from substructures such as the left main coronary artery (LMCA) had more robust radiomic and dosiomic features against perturbation.

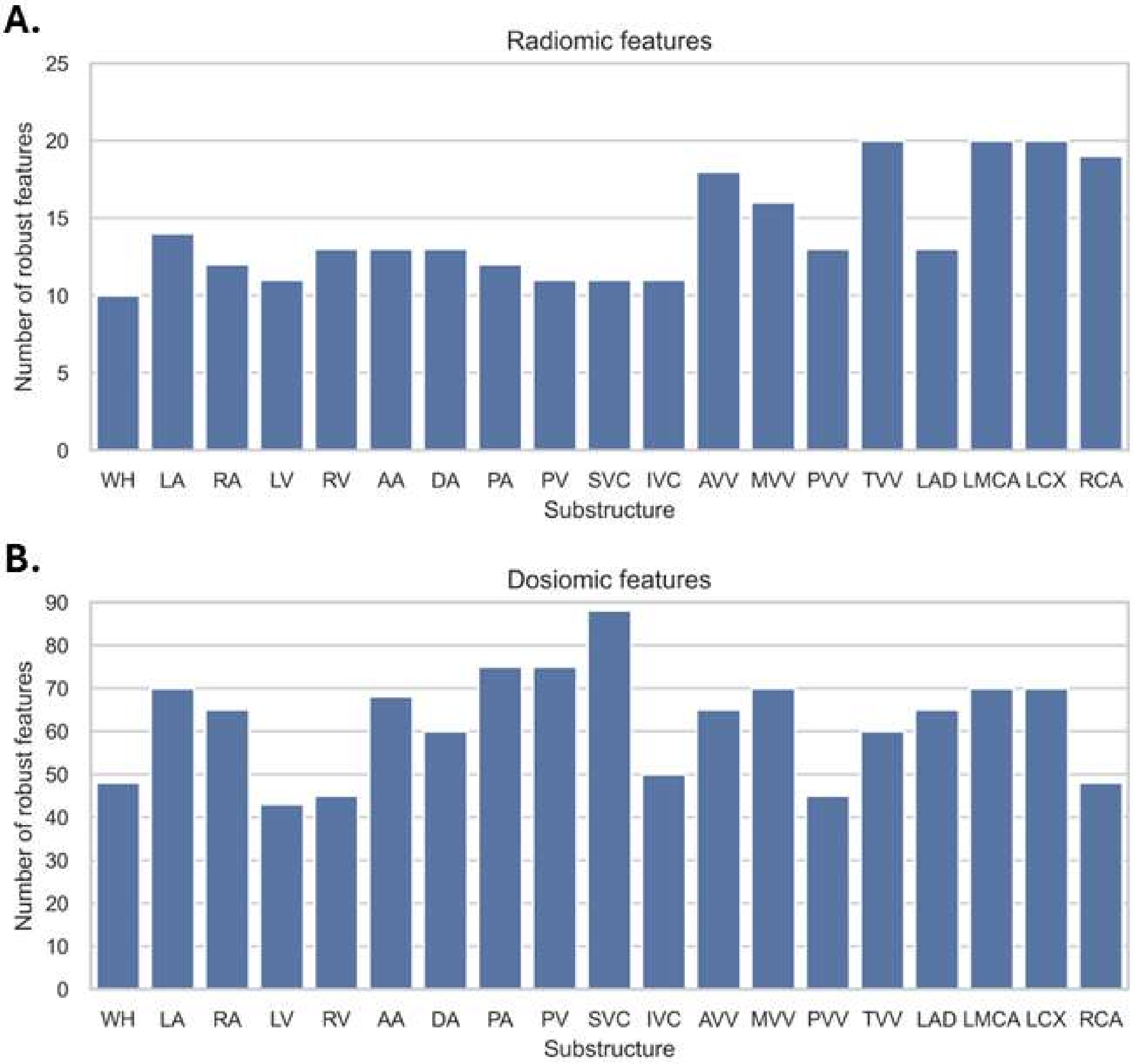


**Fig. A1**. Number of robust radiomic and dosiomic features per substructure. Abbreviations: WH, whole heart; LA, left atrium; RA, right atrium; LV, left ventricle; AA, ascending aorta; DA, descending aorta; PA, pulmonary artery; PV, pulmonary vein; SVC, superior vena cava; IVC, inferior vena cava; AVV, aortic valve; MVV, mitral valve; PVV, pulmonary valve; TVV, tricuspid valve; LAD, left anterior descending coronary artery; LMCA, left main coronary artery; LCX, left circumflex coronary artery; and RCA, right coronary artery.

**Appendix C. Clinical characteristics of photon and proton patients in the retrospective dataset**

| | Value or No. of Patients (%) | | |
|---|---|---|---|
| | Photon (n=98) | Proton (n=62) | *P* Value |
| **Age, years, mean (SD)** | 64.3 (9.1) | 65.3 (8.6) | 0.325 |
| **Sex** | | | 1.000 |
| **Female** | 48 (49) | 30 (48) | |
| **Male** | 50 (51) | 32 (52) | |
| **Race** | | | 1.000 |
| **White** | 89 (91) | 56 (90) | |
| **Other** | 9 (9) | 6 (10) | |
| **Tumor stage** | | | 1.000 |
| **≤IIIA** | 45 (46) | 29 (47) | |
| **≥IIIB** | 53 (54) | 33 (53) | |
| **Tumor histology** | | | 0.158 |
| **Adeno** | 56 (57) | 35 (54) | |
| **Squamous** | 35 (36) | 17 (27) | |
| **Other** | 7 (7) | 10 (16) | |
| **Tumor location** | | | 0.918 |
| **Left/Mediastinal** | 39 (40) | 26 (42) | |
| **Right** | 59 (60) | 36 (58) | |
| **Smoking status** | | | 0.024 |
| **Never** | 10 (10) | 4 (6) | |
| **Former** | 74 (76) | 38 (61) | |
| **Active** | 14 (14) | 20 (32) | |
| **Pre-existing heart disease** | | | 0.716 |
| **No** | 81 (83) | 49 (79) | |
| **Yes** | 17 (17) | 13 (21) | |
| **KPS** | | | 0.861 |
| **<80** | 10 (10) | 5 (8) | |
| **≥80** | 88 (90) | 57 (92) | |
| **hs-cTnT baseline, ng/L, mean (SD)** | 7.2 (8.0) | 9.1 (11.5) | 0.743 |
| **Prescribed dose, Gy, mean (SD)** | 70.3 (4.2) | 70.8 (5.1) | 0.359 |
| **hs-cTnT elevation** | | | 0.138 |
| **No** | 62 (63) | 47 (76) | |
| **Yes** | 36 (37) | 15 (24) | |

Continuous variables are reported as mean (standard deviation) and categorical variables are reported as frequency (percentage). *P* values are from Wilcoxon rank sum test for continuous values and Pearson's Chi-square test for categorical variables.

Abbreviations: KPS, Karnofsky Performance Status score; hs-cTnT, high-sensitivity cardiac troponin T.

**Appendix D. Clinical characteristics of photon and proton patients in the prospective dataset**

| | **Value or No. of Patients (%)** | | |
|---|---|---|---|
| | Photon (n=31) | Proton (n=26) | *P* Value |
| **Age, years, mean (SD)** | 63.9 (9.7) | 72.5 (8.2) | 0.001 |
| **Sex** | | | 0.436 |
| **Female** | 16 (52) | 17 (65) | |
| **Male** | 15 (48) | 9 (35) | |
| **Race** | | | 0.659 |
| **White** | 25 (81) | 23 (88) | |
| **Other** | 6 (19) | 3 (12) | |
| **Tumor stage** | | | 0.770 |
| **≤IIIA** | 18 (58) | 17 (65) | |
| **≥IIIB** | 13 (42) | 9 (35) | |
| **Tumor histology** | | | 0.037 |
| **Adeno** | 15 (49) | 19 (73) | |
| **Squamous** | 10 (32) | 7 (27) | |
| **Other** | 6 (19) | 0 (0) | |
| **Tumor location** | | | 0.766 |
| **Left/Mediastinal** | 12 (39) | 12 (46) | |
| **Right** | 19 (61) | 14 (54) | |
| **Smoking status** | | | 0.208 |
| **Never** | 6 (19) | 7 (27) | |
| **Former** | 23 (74) | 14 (54) | |
| **Active** | 2 (7) | 5 (19) | |
| **Pre-existing heart disease** | | | 0.944 |
| **No** | 24 (77) | 19 (73) | |
| **Yes** | 7 (23) | 7 (27) | |
| **KPS** | | | 0.508 |
| **<80** | 2 (6) | 4 (15) | |
| **≥80** | 29 (94) | 22 (85) | |
| **hs-cTnT baseline, ng/L, mean (SD)** | 9.8 (8.3) | 12.9 (10.3) | 0.152 |
| **Prescribed dose, Gy, mean (SD)** | 66.7 (3.0) | 66.6 (3.4) | 0.359 |
| **hs-cTnT elevation** | | | 1.000 |
| **No** | 22 (71) | 18 (69) | |
| **Yes** | 9 (29) | 8 (31) | |

Continuous variables are reported as mean (standard deviation) and categorical variables are reported as frequency (percentage). *P* values are from Wilcoxon rank sum test for continuous values and Pearson's Chi-square test for categorical variables.

Abbreviations: KPS, Karnofsky Performance Status score; hs-cTnT, high-sensitivity cardiac troponin T.

**Appendix E. Comparison of whole heart (WH)-based and left anterior descending coronary artery (LAD)-based dose-volume histogram (DVH) variables between photon and proton patients in the retrospective and prospective cohorts**

| DVH Variables | Retrospective (n=160) | | | Prospective (n=57) | | |
|---|---|---|---|---|---|---|
| | Photon (n=98) | Proton (n=62) | *P* Value | Photon (n=31) | Proton (n=26) | *P* Value |
| WH V20Gy, % | 25.0 (20.3) | 16.4 (13.6) | 0.023 | 13.6 (13.8) | 7.3 (7.1) | 0.183 |
| WH D60.0, Gy | 6.5 (6.5) | 0.7 (2.9) | <0.0001 | 3.5 (3.9) | 0.3 (0.8) | <0.0001 |
| LAD D1.0cc, Gy | 30.2 (22.5) | 15.4 (19.3) | <0.0001 | 15.7 (17.5) | 11.1 (15.0) | 0.595 |
| LAD V20Gy, % | 34.3 (33.8) | 16.6 (28.2) | 0.0001 | 16.2 (24.9) | 9.0(20.2) | 0.553 |

*P* values were calculated with the Wilcoxon rank-sum test, with a significance level of 0.00015 after Bonferroni correction for multiple comparisons. Numbers were reported as mean (standard deviation).

**Appendix F. Comparison of preselected dose-volume histogram (DVH) variables in patients with and without hs-cTnT elevation in the retrospective and prospective cohorts. All *P* values are from Wilcoxon rank sum tests. Doses are indicated in Gy and dose volumes are in %.**

| Variables | Retrospective (n=160) | | | | Prospective (n=57) | | | | *P** (retrospective vs. prospective) |
|---|---|---|---|---|---|---|---|---|---|
| | All (n=160) | Elevated (n=51) | Non-elevated (n=109) | *p* | All (n=57) | Elevated (n=17) | Non-elevated (n=40) | *p* | |
| WH_V20Gy | 21.7 (18.5) | 28.0 (21.6) | 18.7 (16.0) | 0.016 | 10.7 (11.7) | 15.0 (14.1) | 8.6(9.4) | 0.101 | <0.0001 |
| WH_D60.0 | 4.2 (6.1) | 6.5 (7.8) | 3.2 (4.7) | 0.004 | 2.0 (3.3) | 2.7 (4.4) | 1.4 (2.1) | 0.820 | 0.381 |
| LA_V20Gy | 41.45 (29.45) | 47.47 (30.08) | 39.23 (28.90) | 0.146 | 25.81 (23.06) | 33.14 (27.21) | 22.70 (20.26) | 0.209 | 0.0006 |
| LA_V30Gy | 33.60 (27.04) | 38.40 (27.20) | 31.84 (26.77) | 0.177 | 18.13 (18.44) | 23.82 (20.77) | 15.70 (16.78) | 0.216 | 0.0002 |
| RA_Std | 10.32 (8.48) | 8.20 (7.73) | 11.10 (8.61) | 0.068 | 6.65 (6.48) | 7.39 (6.56) | 6.33 (6.42) | 0.613 | 0.009 |
| RA_Max | 45.02 (29.87) | 38.83 (28.95) | 47.29 (29.88) | 0.091 | 33.50 (26.28) | 38.41 (27.27) | 31.42 (25.56) | 0.464 | 0.003 |
| LV_Max | 30.00 (26.68) | 41.78 (25.86) | 25.67 (25.64) | 0.0005 | 16.97 (18.89) | 21.56 (19.79) | 15.02 (18.15) | 0.279 | 0.002 |
| LV_D0.1cc | 27.47 (26.00) | 39.05 (25.77) | 23.21 (24.75) | 0.0005 | 15.64 (18.29) | 20.27 (19.22) | 13.68 (17.51) | 0.253 | 0.003 |
| RV_V40Gy | 3.74 (7.91) | 4.92 (6.92) | 3.31 (8.20) | 0.002 | 0.95 (3.72) | 0.76 (2.62) | 1.04 (4.09) | 0.862 | 0.0003 |
| RV_Max | 38.84 (23.62) | 47.91 (22.06) | 35.51 (23.30) | 0.002 | 22.97 (18.66) | 25.12 (19.31) | 22.06 (18.30) | 0.74 | <0.0001 |
| AA_Std | 22.57 (6.70) | 21.05 (7.87) | 23.13 (6.12) | 0.159 | 18.31 (6.08) | 17.53 (7.76) | 18.63 (5.17) | 0.701 | <0.0001 |
| AA_Min | 3.31 (4.80) | 4.53 (6.14) | 2.87 (4.11) | 0.231 | 2.91 (6.15) | 2.03 (2.04) | 3.28 (7.19) | 0.754 | 0.322 |
| DA_D40.0 | 23.20 (23.51) | 30.43 (25.10) | 20.55 (22.32) | 0.010 | 18.58 (18.61) | 18.20 (18.58) | 18.75 (18.63) | 0.875 | 0.402 |
| DA_V10Gy | 43.46 (21.16) | 50.48 (19.25) | 40.88 (21.25) | 0.013 | 40.43 (19.40) | 42.34 (23.63) | 39.62 (17.22) | 0.485 | 0.554 |
| PA_Std | 20.48 (7.22) | 18.07 (6.66) | 21.36 (7.22) | 0.006 | 19.43 (5.90) | 18.90 (6.01) | 19.65 (5.84) | 0.65 | 0.281 |
| PA_V10Gy | 86.03 (20.60) | 90.85 (16.92) | 84.26 (21.52) | 0.007 | 76.67 (24.90) | 72.61 (28.21) | 78.39 (23.14) | 0.577 | 0.006 |
| PV_D40.0 | 41.54 (26.98) | 50.11 (24.72) | 38.40 (27.09) | 0.013 | 32.67 (23.59) | 32.31 (24.65) | 32.82 (23.13) | 0.889 | 0.02 |
| PV_D60.0 | 29.66 (25.30) | 36.34 (25.21) | 27.21 (24.89) | 0.018 | 21.29 (20.42) | 19.00 (20.23) | 22.27 (20.42) | 0.507 | 0.027 |
| SVC_V70Gy | 36.38 (36.14) | 22.20 (28.44) | 41.59 (37.25) | 0.005 | 2.37 (9.74) | 3.08 (10.41) | 2.07 (9.43) | 0.212 | <0.0001 |

| | | | | | | | | | |
|---|---|---|---|---|---|---|---|---|---|
| SVC_D70.0 | 51.21 (25.92) | 42.78 (28.13) | 54.30 (24.33) | 0.007 | 37.94 (23.59) | 32.73 (23.23) | 40.16 (23.40) | 0.346 | <0.0001 |
| IVC_D5.0 | 11.59 (19.05) | 12.39 (19.82) | 11.30 (18.75) | 0.335 | 8.79 (17.13) | 15.13 (21.20) | 6.09 (14.25) | 0.045 | 0.31 |
| IVC_Max | 17.14 (23.72) | 18.24 (25.75) | 16.73 (22.92) | 0.369 | 11.62 (19.53) | 19.28 (21.94) | 8.36 (17.42) | 0.012 | 0.229 |
| AVV_Min | 7.00 (9.52) | 9.92 (11.98) | 5.93 (8.17) | 0.154 | 3.82 (6.54) | 4.72 (6.37) | 3.44 (6.58) | 0.565 | 0.09 |
| AVV_V20Gy | 38.88 (40.70) | 46.44 (43.95) | 36.10 (39.07) | 0.171 | 17.65 (30.65) | 28.66 (35.99) | 12.97 (26.74) | 0.257 | <0.0001 |
| MVV_D1.0cc | 24.00 (24.13) | 32.82 (24.59) | 20.76 (23.13) | 0.003 | 11.31 (14.15) | 12.71 (11.98) | 10.72 (14.93) | 0.346 | 0.0005 |
| MVV_V20Gy | 17.95 (29.79) | 28.90 (34.67) | 13.93 (26.67) | 0.004 | 5.47 (15.47) | 8.66 (19.59) | 4.12 (13.10) | 0.524 | 0.0004 |
| PVV_D5.0cc | 26.92 (21.68) | 34.63 (20.55) | 24.08 (21.39) | 0.005 | 17.05 (16.43) | 15.39 (15.74) | 17.76 (16.67) | 0.542 | 0.007 |
| PVV_D60.0$_{60.0}$ | 27.01 (21.58) | 34.28 (20.37) | 24.35 (21.40) | 0.007 | 16.43 (16.38) | 15.67 (16.36) | 16.76 (16.37) | 0.65 | 0.003 |
| TVV_V20Gy | 14.65 (27.20) | 22.29 (31.94) | 11.82 (24.62) | <0.0001 | 3.95 (15.56) | 10.65 (26.60) | 1.10 (4.13) | 0.166 | 0.004 |
| TVV_V40Gy | 3.66 (14.04) | 4.64 (14.44) | 3.29 (13.87) | 0.0001 | 0.02 (0.18) | 0.08 (0.32) | 0.00 (0.00) | 0.485 | 0.066 |
| LAD_D1.0cc | 19.4 (21.2) | 31.2 (22.6) | 13.8 (18.1) | <0.0001 | 12.44 (15.97) | 15.69 (17.54) | 11.06 (15.04) | 0.595 | 0.042 |
| LAD_V20Gy | 21.4 (30.8) | 38.7 (36.9) | 13.2 (23.5) | <0.0001 | 11.13 (21.94) | 16.15 (24.93) | 9.00 (20.17) | 0.553 | 0.014 |
| LMCA_D60.0 | 28.02 (22.86) | 39.56 (21.82) | 23.78 (21.74) | 0.0003 | 14.89 (15.26) | 16.26 (18.92) | 14.30 (13.36) | 0.638 | 0.0004 |
| LMCA_D50.0 | 29.60 (23.31) | 41.45 (22.01) | 25.24 (22.24) | 0.0004 | 15.86 (15.70) | 17.03 (19.30) | 15.36 (13.86) | 0.625 | 0.0003 |
| LCX_V20Gy | 24.77 (34.58) | 39.75 (39.38) | 19.27 (30.86) | 0.146 | 13.34 (23.90) | 15.35 (27.54) | 12.48 (22.12) | 0.807 | 0.022 |
| LCX_D0.1cc | 31.56 (26.96) | 43.95 (25.95) | 27.01 (25.87) | 0.228 | 20.66 (19.56) | 21.39 (21.67) | 20.34 (18.59) | 0.889 | 0.012 |
| RCA_V10Gy | 38.66 (42.93) | 45.89 (45.54) | 36.00 (41.62) | <0.0001 | 21.74 (35.32) | 35.01 (40.74) | 16.10 (31.08) | 0.197 | 0.019 |
| RCA_D5.0cc | 6.56 (11.15) | 8.86 (12.77) | 5.72 (10.36) | 0.0001 | 2.66 (4.99) | 3.69 (6.89) | 2.22 (3.83) | 0.669 | 0.195 |

**P* values in this column indicate the overall comparison between the retrospective and prospective cohorts.

Abbreviations: WH, whole heart; LA, left atrium; RA, right atrium; LV, left ventricle; AA, ascending aorta; DA, descending aorta; PA, pulmonary artery; PV, pulmonary vein; SVC, superior vena cava; IVC, inferior vena cava; AVV, aortic valve; MVV, mitral valve; PVV, pulmonary valve; TVV, tricuspid valve; LAD, left anterior descending coronary artery; LMCA, left main coronary artery; LCX, left circumflex coronary artery; and RCA, right coronary artery.

**Appendix G. Comparison of preselected radiomic variables within patients with and without hs-cTnT elevation in the retrospective and prospective cohorts. All *P* values are from Wilcoxon rank sum tests.**

| Variables | Retrospective (n=160) | | | | Prospective (n=57) | | | | *P** (retrospective vs. prospective) |
|---|---|---|---|---|---|---|---|---|---|
| | All (n=160) | Elevated (n=109) | Non-elevated (n=51) | *p* | All (n=57) | Elevated (n=17) | Non-elevated (n=40) | *p* | |
| WH_CT_shape_Maximum2DDiameterSlice | 135.97 (16.37) | 140.58 (14.83) | 134.28 (16.59) | 0.018 | 133.54 (14.33) | 131.74 (15.27) | 134.30 (13.84) | 0.565 | 0.423 |
| WH_CT_shape_Maximum2DDiameterColumn | 139.99 (14.40) | 141.92 (13.51) | 139.28 (14.64) | 0.168 | 137.74 (12.65) | 134.72 (14.01) | 139.02 (11.79) | 0.346 | 0.399 |
| LA_CT_firstorder_Median | 36.34 (3.75) | 35.65 (3.40) | 36.60 (3.83) | 0.057 | 31.61 (6.79) | 30.65 (2.11) | 32.02 (7.95) | 0.303 | <0.0001 |
| LA_CT_firstorder_RootMeanSquared | 37.00 (4.02) | 36.41 (3.78) | 37.21 (4.09) | 0.119 | 31.50 (7.48) | 30.11 (1.96) | 32.09 (8.77) | 0.229 | <0.0001 |
| RA_CT_shape_LeastAxisLength | 35.09 (5.18) | 36.46 (5.69) | 34.58 (4.89) | 0.087 | 34.35 (5.52) | 33.67 (5.48) | 34.63 (5.51) | 0.875 | 0.534 |
| RA_CT_shape_MeshVolume | 7.22E+04 (2.32E+04) | 7.79E+04 (2.66E+04) | 7.02E+04 (2.14E+04) | 0.148 | 7.10E+04 (2.12E+04) | 6.79E+04 (1.95E+04) | 7.23E+04 (2.18E+04) | 0.613 | 0.986 |
| LV_CT_shape_Flatness | 0.63 (0.05) | 0.64 (0.06) | 0.62 (0.05) | 0.035 | 0.64 (0.06) | 0.65 (0.05) | 0.63 (0.06) | 0.364 | 0.349 |
| LV_CT_shape_LeastAxisLength | 52.13 (5.66) | 52.92 (5.28) | 51.84 (5.77) | 0.184 | 52.42 (4.88) | 51.69 (5.07) | 52.73 (4.76) | 0.78 | 0.649 |
| RV_CT_shape_Maximum2DDiameterSlice | 77.26 (9.25) | 79.15 (9.78) | 76.57 (8.95) | 0.134 | 77.74 (10.46) | 77.09 (10.06) | 78.02 (10.62) | 0.734 | 0.897 |
| RV_CT_shape_MinorAxisLength | 62.43 (6.88) | 63.83 (6.97) | 61.92 (6.77) | 0.155 | 62.29 (7.64) | 61.42 (8.26) | 62.66 (7.32) | 0.374 | 0.889 |
| AA_CT_shape_Maximum2DDiameterSlice | 96.52 (12.70) | 99.88 (11.93) | 95.29 (12.75) | 0.069 | 94.66 (10.51) | 93.63 (13.35) | 95.10 (9.00) | 0.519 | 0.401 |
| AA_CT_shape_Maximum2DDiameterRow | 117.50 (13.41) | 120.15 (13.21) | 116.53 (13.35) | 0.083 | 118.33 (11.82) | 115.15 (11.84) | 119.68 (11.54) | 0.129 | 0.547 |
| DA_CT_shape_Maximum2DDiameterSlice | 38.79 (7.01) | 40.44 (5.91) | 38.18 (7.28) | 0.021 | 37.84 (4.50) | 38.63 (4.79) | 37.50 (4.33) | 0.276 | 0.63 |
| DA_CT_shape_MinorAxisLength | 37.00 (9.06) | 38.45 (8.42) | 36.47 (9.23) | 0.062 | 36.17 (6.74) | 37.77 (6.89) | 35.48 (6.55) | 0.185 | 0.914 |
| PA_CT_shape_MajorAxisLength | 84.67 (11.33) | 88.28 (10.66) | 83.34 (11.28) | 0.018 | 84.45 (10.24) | 82.97 (9.61) | 85.08 (10.43) | 0.393 | 0.926 |
| PA_CT_shape_Maximum2DDiameterSlice | 95.90 (13.20) | 99.40 (12.86) | 94.61 (13.09) | 0.038 | 95.32 (12.03) | 95.30 (11.55) | 95.33 (12.23) | 0.794 | 0.767 |
| PV_CT_shape_Maximum3DDiameter | 105.59 (12.02) | 108.10 (11.28) | 104.67 (12.16) | 0.118 | 104.61 (10.68) | 107.20 (12.83) | 103.51 (9.40) | 0.485 | 0.384 |
| PV_CT_shape_Maximum2DDiameterSlice | 98.92 (12.65) | 101.26 (12.02) | 98.06 (12.77) | 0.162 | 98.47 (11.16) | 100.66 (13.17) | 97.53 (10.04) | 0.619 | 0.546 |
| SVC_CT_shape_MeshVolume | 1.93E+04 (6.22E+03) | 2.04E+04 (6.28E+03) | 1.89E+04 (6.16E+03) | 0.101 | 1.96E+04 (5.26E+03) | 2.07E+04 (6.07E+03) | 1.91E+04 (4.80E+03) | 0.475 | 0.591 |

| | | | | | | | | | |
|---|---|---|---|---|---|---|---|---|---|
| SVC_CT_shape_VoxelVolume | 1.93E+04 (6.23E+03) | 2.04E+04 (6.29E+03) | 1.90E+04 (6.16E+03) | 0.101 | 1.97E+04 (5.26E+03) | 2.08E+04 (6.08E+03) | 1.92E+04 (4.80E+03) | 0.469 | 0.589 |
| IVC_CT_shape_Maximum2DDiameterRow | 49.56 (6.42) | 47.76 (6.39) | 50.22 (6.31) | 0.066 | 48.54 (7.45) | 46.66 (4.59) | 49.35 (8.25) | 0.236 | 0.149 |
| IVC_CT_shape_Maximum3DDiameter | 52.60 (6.72) | 51.03 (4.74) | 53.17 (7.23) | 0.104 | 50.96 (7.02) | 48.88 (4.57) | 51.84 (7.66) | 0.138 | 0.1 |
| AVV_CT_shape_LeastAxisLength | 24.49 (2.92) | 24.83 (2.70) | 24.36 (2.98) | 0.385 | 24.17 (2.82) | 24.54 (3.00) | 24.02 (2.73) | 0.727 | 0.375 |
| AVV_CT_firstorder_TotalEnergy | 3.79E+07 (1.79E+07) | 3.88E+07 (1.50E+07) | 3.76E+07 (1.89E+07) | 0.409 | 2.72E+07 (1.66E+07) | 3.01E+07 (2.36E+07) | 2.60E+07 (1.23E+07) | 0.903 | <0.0001 |
| MVV_CT_firstorder_Mean | 31.91 (5.04) | 30.56 (4.23) | 32.41 (5.21) | 0.032 | 25.90 (8.14) | 25.39 (4.99) | 26.12 (9.14) | 0.944 | <0.0001 |
| MVV_CT_firstorder_Median | 37.00 (3.60) | 36.12 (3.47) | 37.32 (3.60) | 0.063 | 31.79 (6.30) | 31.82 (3.03) | 31.77 (7.25) | 0.663 | <0.0001 |
| PVV_CT_shape_Maximum2DDiameterSlice | 33.09 (4.51) | 34.21 (4.41) | 32.67 (4.48) | 0.065 | 33.77 (3.97) | 33.36 (3.99) | 33.95 (3.94) | 0.688 | 0.378 |
| PVV_CT_shape_MinorAxisLength | 28.87 (3.91) | 29.75 (3.41) | 28.55 (4.03) | 0.084 | 29.08 (3.49) | 28.94 (3.70) | 29.14 (3.39) | 0.727 | 0.778 |
| TVV_CT_shape_LeastAxisLength | 24.01 (3.40) | 24.78 (3.16) | 23.73 (3.44) | 0.089 | 22.94 (3.72) | 22.65 (4.08) | 23.06 (3.55) | 0.834 | 0.112 |
| TVV_CT_shape_MeshVolume | 3.01E+04 (9.43E+03) | 3.23E+04 (1.04E+04) | 2.93E+04 (8.90E+03) | 0.135 | 2.82E+04 (8.32E+03) | 2.76E+04 (8.20E+03) | 2.85E+04 (8.36E+03) | 0.74 | 0.475 |
| LAD_CT_shape_Maximum2DDiameterColumn | 27.29 (10.55) | 31.27 (13.62) | 25.82 (8.73) | 0.036 | 27.11 (8.64) | 31.99 (10.80) | 25.03 (6.51) | 0.023 | 0.51 |
| LAD_CT_shape_Maximum2DDiameterSlice | 36.80 (11.90) | 39.83 (11.41) | 35.69 (11.88) | 0.036 | 36.44 (10.39) | 37.69 (12.28) | 35.90 (9.43) | 0.951 | 0.969 |
| LMCA_CT_firstorder_RootMeanSquared | 45.54 (24.72) | 48.87 (22.96) | 44.31 (25.22) | 0.095 | 42.85 (19.13) | 48.82 (18.39) | 40.31 (18.87) | 0.075 | 0.657 |
| LMCA_CT_firstorder_TotalEnergy | 4.57E+06 (1.11E+07) | 4.58E+06 (6.62E+06) | 4.56E+06 (1.23E+07) | 0.314 | 3.73E+06 (4.95E+06) | 5.80E+06 (6.57E+06) | 2.85E+06 (3.74E+06) | 0.043 | 0.978 |
| LCX_CT_shape_Flatness | 0.20 (0.06) | 0.21 (0.04) | 0.20 (0.07) | 0.021 | 0.22 (0.05) | 0.22 (0.06) | 0.21 (0.04) | 0.986 | 0.003 |
| LCX_CT_glrlm_GrayLevelNonUniformity | 596.92 (242.55) | 674.65 (313.16) | 568.36 (203.41) | 0.026 | 478.18 (156.99) | 470.37 (159.00) | 481.50 (156.01) | 0.958 | 0.0003 |
| RCA_CT_shape_Maximum2DDiameterSlice | 21.59 (6.28) | 23.19 (6.37) | 21.01 (6.14) | 0.058 | 21.21 (5.52) | 22.26 (4.59) | 20.77 (5.81) | 0.316 | 0.602 |
| RCA_CT_glrlm_GrayLevelNonUniformity | 466.86 (238.27) | 525.00 (294.14) | 445.49 (210.10) | 0.085 | 403.97 (182.28) | 465.00 (171.49) | 378.03 (180.53) | 0.112 | 0.099 |

**P* values in this column indicate the overall comparisons between retrospective and prospective patients.

Abbreviations: WH, whole heart; LA, left atrium; RA, right atrium; LV, left ventricle; AA, ascending aorta; DA, descending aorta; PA, pulmonary artery; PV, pulmonary vein; SVC, superior vena cava; IVC, inferior vena cava; AVV, aortic valve; MVV, mitral valve; PVV, pulmonary valve; TVV, tricuspid valve; LAD, left anterior descending coronary artery; LMCA, left main coronary artery; LCX, left circumflex coronary artery; and RCA, right coronary artery.

**Appendix H: Comparison of preselected dosiomic variables within patients with and without hs-cTnT elevation in the retrospective and prospective cohorts. All *P* values are from Wilcoxon rank sum tests.**

| Variables | Retrospective (n=160) | | | | Prospective (n=57) | | | | *P** (retrospective vs. prospective) |
|---|---|---|---|---|---|---|---|---|---|
| | All (n=160) | Elevated (n=109) | Non-elevated (n=51) | *p* | All (n=57) | Elevated (n=17) | Non-elevated (n=40) | *p* | |
| WH_RD_shape_Maximum2DDiameterSlice | 135.97 (16.37) | 140.58 (14.83) | 134.28 (16.59) | 0.018 | 133.54 (14.33) | 131.74 (15.27) | 134.30 (13.84) | 0.565 | 0.423 |
| WH_RD_firstorder_TotalEnergy | 3.34E+08 (2.94E+08) | 4.19E+08 (3.19E+08) | 3.02E+08 (2.79E+08) | 0.035 | 1.35E+08 (1.58E+08) | 1.74E+08 (1.44E+08) | 1.19E+08 (1.61E+08) | 0.062 | <0.0001 |
| LA_RD_firstorder_Energy | 8.08E+07 (7.49E+07) | 1.01E+08 (8.62E+07) | 7.33E+07 (6.88E+07) | 0.069 | 3.80E+07 (4.01E+07) | 4.84E+07 (4.36E+07) | 3.35E+07 (3.77E+07) | 0.222 | 0.0002 |
| LA_RD_firstorder_TotalEnergy | 7.71E+07 (7.14E+07) | 9.64E+07 (8.22E+07) | 7.00E+07 (6.56E+07) | 0.069 | 3.62E+07 (3.83E+07) | 4.61E+07 (4.16E+07) | 3.20E+07 (3.60E+07) | 0.222 | 0.0002 |
| RA_RD_ngtdm_Strength | 0.49 (0.59) | 0.32 (0.40) | 0.55 (0.63) | 0.037 | 0.46 (0.62) | 0.33 (0.31) | 0.51 (0.71) | 0.794 | 0.314 |
| RA_RD_glcm_ClusterShade | 697.54 (1843.25) | 68.68 (1386.28) | 928.65 (1934.22) | 0.04 | 542.57 (895.96) | 503.44 (974.09) | 559.19 (860.08) | 0.794 | 0.714 |
| LV_RD_firstorder_Maximum | 28.97 (26.36) | 40.85 (25.73) | 24.61 (25.23) | 0.0003 | 17.35 (19.29) | 22.17 (20.41) | 15.30 (18.42) | 0.257 | 0.006 |
| LV_RD_firstorder_Range | 28.18 (25.85) | 39.63 (25.29) | 23.98 (24.75) | 0.0003 | 16.93 (19.03) | 21.27 (20.06) | 15.09 (18.26) | 0.329 | 0.006 |
| RV_RD_firstorder_Maximum | 37.26 (23.37) | 46.73 (21.99) | 33.78 (22.90) | 0.001 | 23.07 (18.91) | 25.79 (19.73) | 21.91 (18.43) | 0.727 | <0.0001 |
| RV_RD_firstorder_Range | 36.26 (22.90) | 45.35 (22.01) | 32.91 (22.30) | 0.002 | 22.64 (18.59) | 24.91 (19.36) | 21.68 (18.17) | 0.807 | 0.0001 |
| AA_RD_glrlm_RunVariance | 21.62 (28.58) | 24.23 (28.75) | 20.66 (28.46) | 0.024 | 28.93 (36.70) | 39.68 (50.23) | 24.36 (27.87) | 0.74 | 0.314 |
| AA_RD_glrlm_LongRunEmphasis | 41.99 (79.22) | 46.14 (66.20) | 40.46 (83.44) | 0.028 | 60.64 (108.77) | 92.73 (166.74) | 47.00 (66.48) | 0.565 | 0.399 |
| DA_RD_gldm_LargeDependenceHighGrayLevelEmphasis | 6.53E+04 (7.49E+04) | 8.82E+04 (8.25E+04) | 5.69E+04 (7.01E+04) | 0.006 | 4.29E+04 (5.06E+04) | 4.76E+04 (4.66E+04) | 4.09E+04 (5.20E+04) | 0.346 | 0.189 |
| DA_RD_glrlm_RunEntropy | 6.58 (0.44) | 6.73 (0.27) | 6.52 (0.47) | 0.006 | 6.49 (0.57) | 6.46 (0.68) | 6.50 (0.52) | 0.714 | 0.838 |
| PA_RD_firstorder_Energy | 1.86E+08 (9.39E+07) | 2.22E+08 (9.93E+07) | 1.73E+08 (8.82E+07) | 0.004 | 1.16E+08 (6.24E+07) | 1.15E+08 (5.73E+07) | 1.16E+08 (6.45E+07) | 0.917 | <0.0001 |
| PA_RD_firstorder_TotalEnergy | 1.77E+08 (8.96E+07) | 2.12E+08 (9.47E+07) | 1.65E+08 (8.41E+07) | 0.004 | 1.11E+08 (5.96E+07) | 1.10E+08 (5.47E+07) | 1.11E+08 (6.15E+07) | 0.917 | <0.0001 |

| | | | | | | | | | |
|---|---|---|---|---|---|---|---|---|---|
| PV_RD_glcm_ClusterShade | 1.37E+03 (8.18E+03) | -1629.88 (7226.79) | 2.47E+03 (8.23E+03) | 0.009 | 2.35E+03 (5.49E+03) | 2.93E+03 (5.36E+03) | 2.10E+03 (5.53E+03) | 0.485 | 0.342 |
| PV_RD_firstorder_Median | 34.83 (26.76) | 43.03 (25.18) | 31.81 (26.70) | 0.012 | 25.79 (22.27) | 25.36 (22.47) | 25.98 (22.18) | 0.903 | 0.02 |
| SVC_RD_firstorder_Skewness | -0.88 (1.58) | -0.52 (1.23) | -1.02 (1.68) | 0.019 | 0.17 (3.87) | -0.13 (1.80) | 0.30 (4.47) | 0.443 | 0.105 |
| SVC_RD_firstorder_RootMeanSquared | 55.72 (21.58) | 50.38 (23.41) | 57.68 (20.53) | 0.028 | 43.34 (21.42) | 42.02 (19.05) | 43.90 (22.33) | 0.432 | <0.0001 |
| IVC_RD_shape_Maximum2DDiameterRow | 49.56 (6.42) | 47.76 (6.39) | 50.22 (6.31) | 0.066 | 48.54 (7.45) | 46.66 (4.59) | 49.35 (8.25) | 0.236 | 0.149 |
| IVC_RD_shape_Maximum3DDiameter | 52.60 (6.72) | 51.03 (4.74) | 53.17 (7.23) | 0.104 | 50.96 (7.02) | 48.88 (4.57) | 51.84 (7.66) | 0.138 | 0.1 |
| AVV_RD_firstorder_Minimum | 7.33 (9.96) | 10.36 (12.53) | 6.21 (8.57) | 0.158 | 3.71 (6.31) | 4.57 (6.18) | 3.34 (6.34) | 0.589 | 0.069 |
| AVV_RD_firstorder_10Percentile | 10.31 (13.11) | 13.26 (15.06) | 9.23 (12.14) | 0.182 | 5.12 (8.67) | 6.56 (8.21) | 4.51 (8.79) | 0.312 | 0.027 |
| MVV_RD_glszm_ZoneEntropy | 3.16 (1.90) | 3.84 (1.77) | 2.91 (1.88) | 0.002 | 2.31 (1.78) | 2.66 (1.76) | 2.17 (1.76) | 0.283 | 0.003 |
| MVV_RD_firstorder_TotalEnergy | 1.23E+07 (2.36E+07) | 1.89E+07 (2.69E+07) | 9.89E+06 (2.18E+07) | 0.003 | 3.10E+06 (9.19E+06) | 2.78E+06 (4.51E+06) | 3.23E+06 (1.06E+07) | 0.264 | 0.001 |
| PVV_RD_firstorder_Minimum | 16.03 (15.97) | 20.74 (14.22) | 14.30 (16.23) | 0.006 | 9.35 (11.98) | 11.21 (13.60) | 8.56 (11.13) | 0.607 | 0.013 |
| PVV_RD_firstorder_10Percentile | 20.97 (19.14) | 27.05 (17.20) | 18.73 (19.32) | 0.006 | 12.32 (13.74) | 13.02 (14.76) | 12.02 (13.28) | 0.993 | 0.007 |
| TVV_RD_shape_LeastAxisLength | 24.01 (3.40) | 24.78 (3.16) | 23.73 (3.44) | 0.089 | 22.94 (3.72) | 22.65 (4.08) | 23.06 (3.55) | 0.834 | 0.112 |
| TVV_RD_shape_VoxelVolume | 3.01E+04 (9.43E+03) | 3.24E+04 (1.04E+04) | 2.93E+04 (8.91E+03) | 0.135 | 2.83E+04 (8.33E+03) | 2.77E+04 (8.20E+03) | 2.85E+04 (8.36E+03) | 0.74 | 0.477 |
| LAD_RD_glszm_ZoneEntropy | 3.29 (1.75) | 4.09 (1.44) | 2.99 (1.77) | <0.0001 | 2.61 (1.71) | 2.74 (1.76) | 2.56 (1.68) | 0.708 | 0.013 |
| LAD_RD_firstorder_90Percentile | 22.94 (22.73) | 33.81 (22.95) | 18.94 (21.29) | <0.0001 | 14.05 (16.87) | 17.15 (19.07) | 12.74 (15.65) | 0.754 | 0.012 |
| LMCA_RD_firstorder_10Percentile | 22.94 (20.78) | 32.96 (20.65) | 19.25 (19.58) | 0.0002 | 11.70 (13.72) | 13.75 (17.27) | 10.82 (11.79) | 0.848 | 0.001 |
| LMCA_RD_firstorder_Median | 29.47 (23.12) | 41.28 (21.88) | 25.12 (22.01) | 0.0002 | 15.57 (15.75) | 17.00 (19.29) | 14.97 (13.94) | 0.695 | 0.0002 |
| LCX_RD_firstorder_Maximum | 36.24 (27.47) | 48.84 (24.70) | 31.60 (26.97) | 0.0003 | 24.18 (21.86) | 24.29 (23.23) | 24.13 (21.25) | 0.727 | 0.003 |
| LCX_RD_firstorder_90Percentile | 26.86 (25.77) | 38.28 (25.73) | 22.66 (24.48) | 0.0004 | 15.82 (17.21) | 17.76 (18.96) | 14.99 (16.34) | 0.972 | 0.008 |
| RCA_RD_shape_Maximum2DDiameterSlice | 21.59 (6.28) | 23.19 (6.37) | 21.01 (6.14) | 0.058 | 21.21 (5.52) | 22.26 (4.59) | 20.77 (5.81) | 0.316 | 0.602 |

| | | | | | | | | | |
|---|---|---|---|---|---|---|---|---|---|
| RCA_RD_glrlm_LongRunHighGrayLevelEmphasis | 359.88 (579.20) | 393.52 (544.08) | 347.52 (591.10) | 0.135 | 235.40 (380.93) | 355.62 (395.14) | 184.31 (362.86) | 0.01 6 | 0.523 |

*$P$ values in this column indicate the overall comparisons between patients in the retrospective and prospective cohorts.

Abbreviations: WH, whole heart; LA, left atrium; RA, right atrium; LV, left ventricle; AA, ascending aorta; DA, descending aorta; PA, pulmonary artery; PV, pulmonary vein; SVC, superior vena cava; IVC, inferior vena cava; AVV, aortic valve; MVV, mitral valve; PVV, pulmonary valve; TVV, tricuspid valve; LAD, left anterior descending coronary artery; LMCA, left main coronary artery; LCX, left circumflex coronary artery; RCA, right coronary artery.
